# Prevention and Attenuation of COVID-19 by BNT162b2 and mRNA-1273 Vaccines

**DOI:** 10.1101/2021.06.01.21257987

**Authors:** Mark G. Thompson, Jefferey L. Burgess, Allison L. Naleway, Harmony Tyner, Sarang K. Yoon, Jennifer Meece, Lauren E.W. Olsho, Alberto J. Caban-Martinez, Ashley L. Fowlkes, Karen Lutrick, Holly C. Groom, Kayan Dunnigan, Marilyn J Odean, Kurt Hegmann, Elisha Stefanski, Laura J. Edwards, Natasha Schaefer-Solle, Lauren Grant, Katherine Ellingson, Jennifer L. Kuntz, Tnelda Zunie, Matthew S. Thiese, Lynn Ivacic, Meredith G. Wesley, Julie Mayo Lamberte, Xiaoxiao Sun, Michael E. Smith, Andrew L. Phillips, Kimberly D. Groover, Young M. Yoo, Joe K. Gerald, Rachel T. Brown, Meghan K. Herring, Gregory Joseph, Shawn Beitel, Tyler C. Morrill, Josephine Mak, Patrick Rivers, Brandon P. Poe, Brian Lynch, Ying Tao Zhou, Jing Zhang, Anna Kelleher, Yan Li, Monica Dickerson, Erika Hanson, Kyley Guenther, Suxiang Tong, Allen Bateman, Erik Reisdorf, John Barnes, Eduardo Azziz-Baumgartner, Danielle R. Hunt, Melissa L. Arvay, Preeta Kutty, Alicia M. Fry, Manjusha Gaglani

**Affiliations:** CDC COVID-19 Response Team; Mel and Enid Zuckerman College of Public Health, University of Arizona; Kaiser Permanente Northwest Center for Health Research; St. Luke’s; University of Utah; Marshfield Clinic Research Laboratory; Abt Associates, Inc.; Leonard M. Miller School of Medicine, University of Miami; Baylor Scott & White Health; Whiteside Institute for Clinical Research, St. Luke’s; Texas A&M University College of Medicine; Wisconsin State Laboratory of Hygiene

**Author notes:** Corresponding Author: Mark G Thompson, PhD. Disclaimer: The findings and conclusions in this report are those of the authors and do not necessarily represent the official position of the Centers for Disease Control and Prevention.

## Abstract

**BACKGROUND:** Information is limited on messenger RNA (mRNA) BNT162b2 (Pfizer-BioNTech) and mRNA-1273 (Moderna) COVID-19 vaccine effectiveness (VE) in preventing SARS-CoV-2 infection or attenuating disease when administered in real-world conditions.

**METHODS:** Prospective cohorts of 3,975 healthcare personnel, first responders, and other essential and frontline workers completed weekly SARS-CoV-2 testing during December 14 2020—April 10 2021. Self-collected mid-turbinate nasal swabs were tested by qualitative and quantitative reverse-transcription–polymerase-chain-reaction (RT-PCR). VE was calculated as 100%×(1−hazard ratio); adjusted VE was calculated using vaccination propensity weights and adjustments for site, occupation, and local virus circulation.

**RESULTS:** SARS-CoV-2 was detected in 204 (5.1%) participants; 16 were partially (≥14 days post-dose-1 to 13 days after dose-2) or fully (≥14 days post-dose-2) vaccinated, and 156 were unvaccinated; 32 with indeterminate status (<14 days after dose-1) were excluded. Adjusted mRNA VE of full vaccination was 91% (95% confidence interval [CI]=76%–97%) against symptomatic or asymptomatic SARS-CoV-2 infection; VE of partial vaccination was 81% (95% CI=64%-90%). Among partially or fully vaccinated participants with SARS-CoV-2 infection, mean viral RNA load (Log10 copies/mL) was 40% lower (95% CI=16%-57%), the risk of self-reported febrile COVID-19 was 58% lower (Risk Ratio=0.42, 95% CI=0.18-0.98), and 2.3 fewer days (95% CI=0.8-3.7) were spent sick in bed compared to unvaccinated infected participants.

**CONCLUSIONS:** Authorized mRNA vaccines were highly effective among working-age adults in preventing SARS-CoV-2 infections when administered in real-world conditions and attenuated viral RNA load, febrile symptoms, and illness duration among those with breakthrough infection despite vaccination.

---

Messenger RNA (mRNA) BNT162b2 (Pfizer-BioNTech) and mRNA-1273 (Moderna) COVID-19 two-dose vaccines were highly effective in preventing symptomatic COVID-19 in randomized placebo-controlled Phase III efficacy trials ^1,2^. Recently, we reported interim estimates of vaccine effectiveness (VE) in preventing symptomatic and asymptomatic SARS-CoV-2 infection that showed similar benefits for mRNA vaccines administered in real-world conditions ^3^. Less is known about the potentially important secondary benefits of mRNA COVID-19 vaccine, including reductions in severity of disease, viral RNA load, and duration of viral RNA detection.

Using prospective cohorts of healthcare personnel, first responders, and other essential and frontline workers in eight U.S. locations, this report had three aims. First, we estimated VE of partial and full mRNA vaccination in preventing SARS-CoV-2 infections, with adjustments for propensity to be vaccinated and local virus circulation. Second, among participants with laboratory-confirmed SARS-CoV-2 infection, we compared the viral RNA load of participants who were partially or fully vaccinated with those who were unvaccinated. Third, we compared the frequency of febrile symptoms and the duration of COVID-19 among infected participants who were partially or fully vaccinated versus unvaccinated infected participants.

## METHODS

### Study Population

HEROES-RECOVER^i^ is a network of prospective cohorts in eight locations (Phoenix, Tucson, and other areas in Arizona; Miami, Florida; Duluth, Minnesota; Portland, Oregon; Temple, Texas; and Salt Lake City, Utah) that initiated in July 2020 and share a common protocol, described previously and in Supplementary_Appendix_Methods. To minimize potential selection biases, participants were invited to join the cohorts using a sampling strategy stratified by site, sex, age group, and occupation. The present analysis was conducted during December 14, 2020-April 10, 2021. All participants provided written consent and both the RECOVER and HEROES studies were reviewed and approved by the Institutional Review Boards at participating sites or under a reliance arrangement.^ii^

### Measures

Socio-demographic and health characteristics were self-reported by electronic surveys at enrollment. Each month, participants reported potential SARS-CoV-2 exposure time and their use of face masks and other employer recommended personal protective equipment (PPE) with four measures: hours of close contact (within 3 feet) of others at work (co-workers, customers, patients, or the public) in the past 7 days; percent time using PPE during those hours; hours of close contact with someone suspected or confirmed to have COVID-19 at work, home, or the community in the past 7 days; percent time using PPE during this contact.

Active surveillance for symptoms consistent with COVID-19–like illness (defined as fever, chills, cough, shortness of breath, sore throat, diarrhea, muscle aches, or change in smell or taste) occurred through weekly text messages, e-mails, and direct participant or medical record reports. When COVID-19–like illness was identified, participants self-reported the symptom onset date and completed electronic surveys at the beginning and end of illness to indicate symptoms, temperature, days sick in bed for at least half the day, receipt of medical care, and last day symptomatic or ill (Supplementary_Appendix_Methods). Febrile COVID-19–like illness was defined as fever, feverishness, chills, or measured temperature >38 degrees Celsius.

### Laboratory Methods

Participants self-collected a mid-turbinate nasal swab weekly, regardless of COVID-19–like illness symptoms, and collected an additional nasal swab and saliva specimen at the onset of COVID-19–like illness. Supplies and instructions for participants were standardized across sites. Specimens were shipped weekdays on cold packs and were tested by reverse-transcription-polymerase chain reaction (RT-PCR) assay at Marshfield Clinic Laboratory (Marshfield, Wisconsin). Quantitative RT-PCR was conducted by the Wisconsin State Laboratory of Hygiene (Madison, Wisconsin). SARS-CoV-2 whole genome sequencing was conducted at CDC using previously published protocols^4^ for viruses detected among 22 participants who were ≥7 days post dose-1 at infection (through March 3, 2021) and 3-4 unvaccinated participants (71 total) location-and closest-date-matched, as available. Viral lineages were categorized as variants of concern, variants of interest, or other. We compare the percentage of variants of concern (excluding variants of interest) among participants who are at least partially vaccinated (≥14-days post-dose-1) with those unvaccinated. Further details on laboratory methods are in Supplementary_Appendix_Methods.

### Vaccination Status

COVID-19 vaccination status was documented by self-report in electronic and telephone surveys and through direct upload of vaccination card images. Additionally, data from electronic medical records, occupational health records, or state immunization registries were reviewed at the Minnesota, Oregon, Texas, and Utah sites (Supplementary_Appendix_Methods).

### Statistical Analysis

The primary outcome was time to RT-PCR confirmed SARS-CoV-2 infection in vaccinated versus unvaccinated participants. Secondary outcomes included viral RNA load, febrile illness, and illness duration. VE was estimated for full vaccination (≥14-days after mRNA vaccine dose-2) and partial vaccination (≥14-days post-dose-1 and up to 13-days post-dose-2). Days 1-13 following dose-1 were excluded because vaccination status was considered indeterminate. Hazard ratios were estimated by the Andersen-Gill extension of the Cox proportional hazards model, which accounted for time-varying vaccination status. Unadjusted VE was calculated as 100%×(1−hazard ratio). An adjusted VE model accounted for potential confounding in vaccination status using an inverse propensity of treatment weighting (IPTW) approach ^5^. Generalized boosted regression trees were used to estimate individual propensities to be at least partially vaccinated during each study week conditional on baseline socio-demographic and health characteristics and the most recent reports of virus exposure and PPE use (Table 1 and Table_S2)^6^. Predicted propensities were then used to calculate stabilized IPTW weights. Cox models incorporated these stabilized weights as well as covariates for site and occupation and a daily indicator of local virus circulation (the percentage positive of all SARS-CoV-2 testing for local counties; Figure_S1). A sensitivity analysis removed person-days among participants with possible misclassification of vaccination or infection and at locations when local SARS-CoV-2 circulation fell below 3% as described in Supplementary_Appendix_Methods.

**Table 1.**
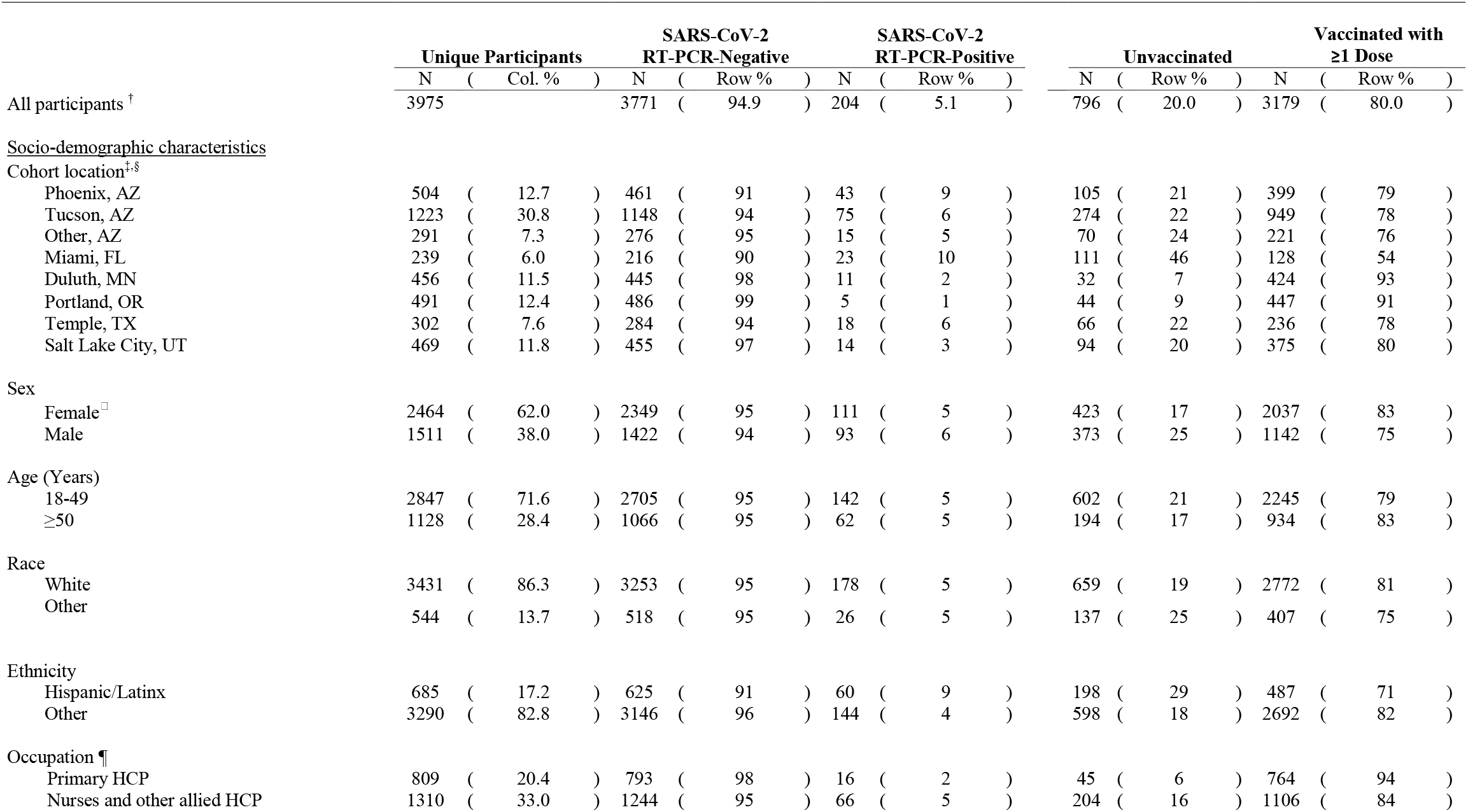
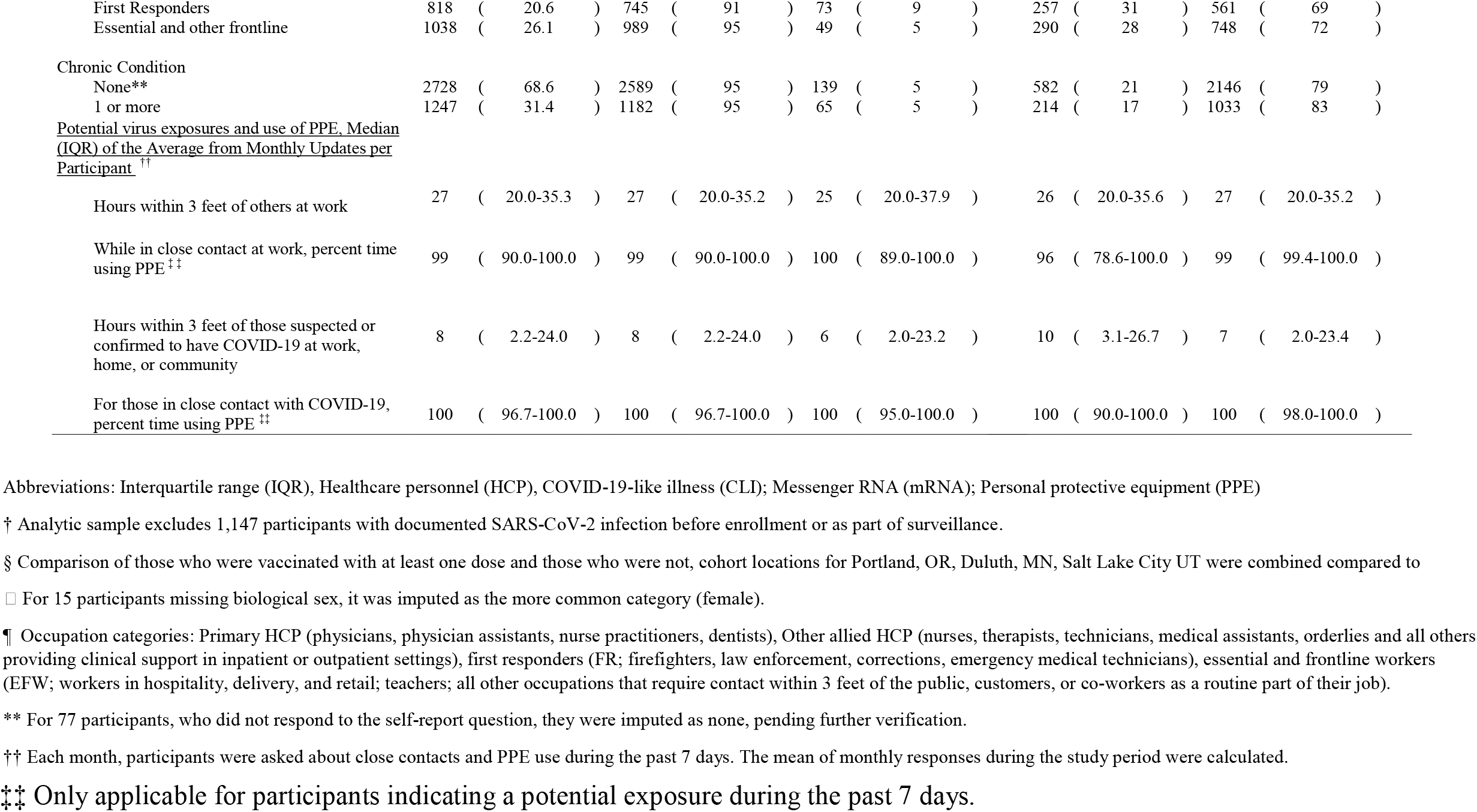
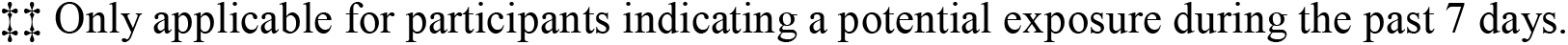
Characteristics of healthcare personnel (HCP), first responders, and other essential and frontline workers in prospective cohorts and percentage with RT-PCR-confirmed SARS-CoV-2 infections and percentage receiving ≥1 dose of messenger RNA COVID vaccine during the study period

Due to the relatively small number of breakthrough infections, for evaluation of possible vaccination attenuation effects, participants with RT-PCR-confirmed infections while partially or fully vaccinated were combined into a single vaccinated group and compared to participants who were unvaccinated when infected. The highest viral RNA load (Log10 copies/mL) measured during RT-PCR-confirmed infections were compared using a Poisson model adjusted for days from symptom onset to specimen collection and for days in transit to the laboratory.

Dichotomous outcomes were compared using binary log-logistic regression to calculate relative risks RR). Comparisons of illness duration outcomes in days were made with Student’s t-test assuming unequal variances. All analyses were conducted with SAS (version 9.4; SAS Institute) and R (version 4.0.2; R Foundation for Statistical Computing).

## RESULTS

### Participant Characteristics

After exclusion of 1,147 participants with laboratory documentation of SARS-CoV-2 infection before the start of the study period, the analytic sample consisted of 3,975 participants; CONSORT diagram is Figure_S2. Approximately one-half of the participants (50.8%) were from the three Arizona study sites (Table 1). The majority of participants were female (62.0%), aged 18–49 years (71.6%), White (86.3%), and non-Hispanic (82.8%) and had no chronic medical conditions (68.6%). Participants included primary healthcare providers (20.4%), such as physicians and other clinical leads, nurses and other allied healthcare personnel (33.0%), first responders (20.6%), and other essential and frontline workers (26.1%). Over the 17-week study period, adherence to weekly surveillance reporting and specimen collection was high (median = 100%; interquartile range = 82%–100%). COVID-19

### COVID-19 Vaccination Status

Most (3,179/3,975, 80.0%) of the participants received at least one dose of an authorized mRNA COVID-19 vaccine by April 10, 2021 (Table 1); 2,686/3,179 (84.5%) received both recommended doses; vaccine products were 66.8% Pfizer-BioNTech, 32.7% Moderna, and 0.6% unspecified mRNA vaccine. Too few participants received Johnson & Johnson/Janssen COVID-19 vaccine (n=39) for comparison with mRNA vaccines, and therefore their person-time was censored at vaccination, and they contributed only unvaccinated person-time. Receipt of at least one vaccine dose was higher among participants in Minnesota or Oregon and participants who were female, aged ≥50 years, White, non-Hispanic, healthcare personnel, or had ≥1 chronic health condition. Mean hours of close contact with people suspected or confirmed to have COVID-19 was lower and the percentage of time using PPE was higher among vaccinated participants (Table 1). Associations with additional covariates included in the vaccination propensity model are presented in Table_S2. Standardized mean differences between vaccinated and unvaccinated were well balanced after propensity weighting with a maximum difference of 0.09 (Figure_S3).

### SARS-CoV-2 RT-PCR-confirmed Infections

SARS-CoV-2 infection was detected by RT-PCR in 204 (5.1%) participants, including 5 fully and 16 partially vaccinated, and 156 unvaccinated; 32 with indeterminate vaccination status (<14 days post-dose-1) were excluded. Of 93 genetically sequenced viruses, 10 were variants of concern (8 B.1.429, 1 B.1.1.7, 1 B.1.427) and 1 was a variant of interest (P.2) (Table_S3). Excluding 12 viruses among those with indeterminate vaccination, 10 viruses were detected among partially or fully vaccinated participants; of these, 3 (30.0%) were variants of concern (all B.1.429) compared to 7/70 (10.0%) among unvaccinated participants.

RT-PCR-confirmed infection was higher among participants in Arizona, Florida, and Texas or and those who were males, Hispanics, or first responders (Table 1); however, infection outcome did not differ by reported hours of potential virus exposure and PPE use. The majority of RT-PCR-confirmed infections had COVID-19–like illness symptoms before or within 1 day of specimen collection (74.0%) or within 2-14 days after collection (13.2%); the remainder had other symptoms (2.0%) or were asymptomatic within 14 days before and after specimen collection (10.8%). Only 26.0% of PCR-confirmed infections were medically attended, including two hospitalizations among unvaccinated participants; no deaths were reported.

### mRNA COVID-19 Vaccine Effectiveness

During the 17-week study period, 3,975 participants contributed a median of 19 (Interquartile Range [IQR]=8-41) unvaccinated days per participant (127,971 total); 156 RT-PCR-confirmed SARS-CoV-2 infections were identified. While partially vaccinated, 3,001 participants contributed a median of 22 (IQR=21-28) days (81,168 total), during which 11 RT-PCR-confirmed infections were identified. While fully vaccinated, 2,510 participants contributed a median of 69 (IQR=53-81) days (161,613 total) during which 5 RT-PCR-confirmed infections were identified. Results of vaccination propensity weight calculations are shown in Figure_S3. Estimated adjusted VE of full vaccination against RT-PCR confirmed infection was 91% (95% confidence interval [CI] = 76%–97%); VE of partial vaccination against RT-PCR confirmed infection was 81% (95% CI=64%–90%) (Table 2). Secondary VE estimates by mRNA vaccine product and age group are available in Table 2. VE point estimates were unchanged in a sensitivity analysis that excluded periods of low local virus circulation (Table_S4).

**Table 2.** Contributing participants, person-days, and the number of RT-PCR confirmed SARS-CoV-2 infections by vaccination status with estimates of messenger RNA (mRNA) vaccine effectiveness (VE) for partial and full vaccination in preventing infection among 3,975 healthcare personnel, first responders, and essential and frontline workers

|  | Contributing<br>Participants<sup>*</sup> | Total Person-Days | Median (IQR)<br>Days | SARS-CoV-2<br>Infections<sup>*</sup> | Unadjusted VE<br>% ( 95% CI ) | Adjusted VE<sup>†</sup><br>% ( 95% CI ) |
| --- | --- | --- | --- | --- | --- | --- |
| <u>mRNA COVID-19 vaccination status</u> |  |  |  |  |  |  |
| Unvaccinated | 3,964 | 127,971 | 19 (8 - 41) | 156 |  |  |
| Partially vaccinated (≥14-days post dose-1 to day 13 post dose-2) | 3,001 | 81,168 | 22 (21 - 28) | 11 | 86 ( 74 - 93 ) | 81 ( 64 - 90 ) |
| Fully vaccinated (≥14-days post dose-2) | 2,510 | 161,613 | 69 (53 - 81) | 5 | 92 ( 80 - 97 ) | 91 ( 76 - 97 ) |
| <u>Pfizer-BioNTech</u> |  |  |  |  |  |  |
| Unvaccinated | 3,964 | 127,971 | 19 (8 - 41) | 156 |  |  |
| Partially vaccinated (≥14-days post dose-1 to day 13 post dose-2) | 2,005 | 49,516 | 21 (21 - 22) | 8 | 85 ( 69 - 93 ) | 80 ( 60 - 90 ) |
| Fully vaccinated (≥14-days post dose-2) | 1,731 | 120,653 | 77 (64 - 82) | 3 | 94 ( 82 - 98 ) | 93 ( 78 - 98 ) |
| <u>Moderna</u> |  |  |  |  |  |  |
| Unvaccinated | 3,964 | 127,971 | 19 (8 - 41) | 156 |  |  |
| Partially vaccinated (≥14-days post dose-1 to day 13 post dose-2) | 982 | 31,231 | 28 (28 - 31) | 3 | 88 ( 61 - 96 ) | 83 ( 40 - 95 ) |
| Fully vaccinated (≥14-days post dose-2) | 770 | 40,394 | 58 (44 - 66) | 2 | 84 ( 31 - 96 ) | 82 ( 20 - 96 ) |
| <u>Age < 50</u> |  |  |  |  |  |  |
| Unvaccinated | 2,838 | 90,768 | 18 (8 - 42) | 107 |  |  |
| Partially vaccinated (≥14-days post dose-1 to day 13 post dose-2) | 2,116 | 57,064 | 22 (21 - 28) | 8 | 87 ( 72 - 94 ) | 81 ( 59 - 91 ) |
| Fully vaccinated (≥14-days post dose-2) | 1,760 | 114,676 | 72 (55 - 81) | 4 | 91 ( 75 - 97 ) | 90 ( 69 - 97 ) |
| <u>Age ≥ 50</u> |  |  |  |  |  |  |
| Unvaccinated | 1,126 | 37,203 | 21 (9 - 40) | 49 |  |  |
| Partially vaccinated (≥14-days post dose-1 to day 13 post dose-2) | 885 | 24,104 | 22 (21 - 28) | 3 | 84 ( 46 - 95 ) | 78 ( 28 - 93 ) |
| Fully vaccinated (≥14-days post dose-2) | 750 | 46,937 | 68 (50 - 80) | 1 | 95 ( 59 - 99 ) | 94 ( 51 - 99 ) |

Abbreviations: Messenger RNA (mRNA), Vaccine effectiveness (VE), Interquartile range (IQR)
\* Contributing participants in vaccination categories do not equal the number of vaccination doses because participants must be in active surveillance and have met the vaccination criteria. Person-days during days 1-13 following dose-1 and the 32 associated RT-PCR-confirmed infections are excluded from this model because immunity was considered indeterminate.
† Adjusted VE is inversely weighted for propensity to be vaccinated with doubly robust adjustment for local virus circulation, study location, and occupation (Supplementary\_Appendix\_Methods)

### Comparisons Between Partially or Fully Vaccinated and Unvaccinated RT-PCR-Confirmed SARS-CoV-2 Infected Participants

Characteristics of 16 participants who were partially or fully vaccinated at SARS-CoV-2 infection and 156 participants who were unvaccinated at the time of infection are listed in Table_S5. The percentage infected while partially or fully vaccinated was higher among participants in Arizona, Minnesota, and Utah and among healthcare personnel; otherwise, there were no other differences by socio-demographic, health, virus exposure, or PPE use (Table_S5).

### Vaccine Attenuation of Viral RNA Load

Mean viral RNA load measured for 155 participants was not associated with participant characteristics, except for somewhat lower viral RNA load among first responders (Table_S6). The mean viral RNA load (Log10 copies/mL) detected for SARS-CoV-2 infections was 3.8 among unvaccinated and 2.3 among partially or fully vaccinated participants; in an adjusted model, this represented 40.2% (95% CI=16.3%-57.3%) lower viral RNA load after at least partial vaccination (Table 3). Examining viral RNA only among those vaccinated, mean viral RNA load detected declined after receipt of dose-1 (Figure_S4).

**Table 3.**
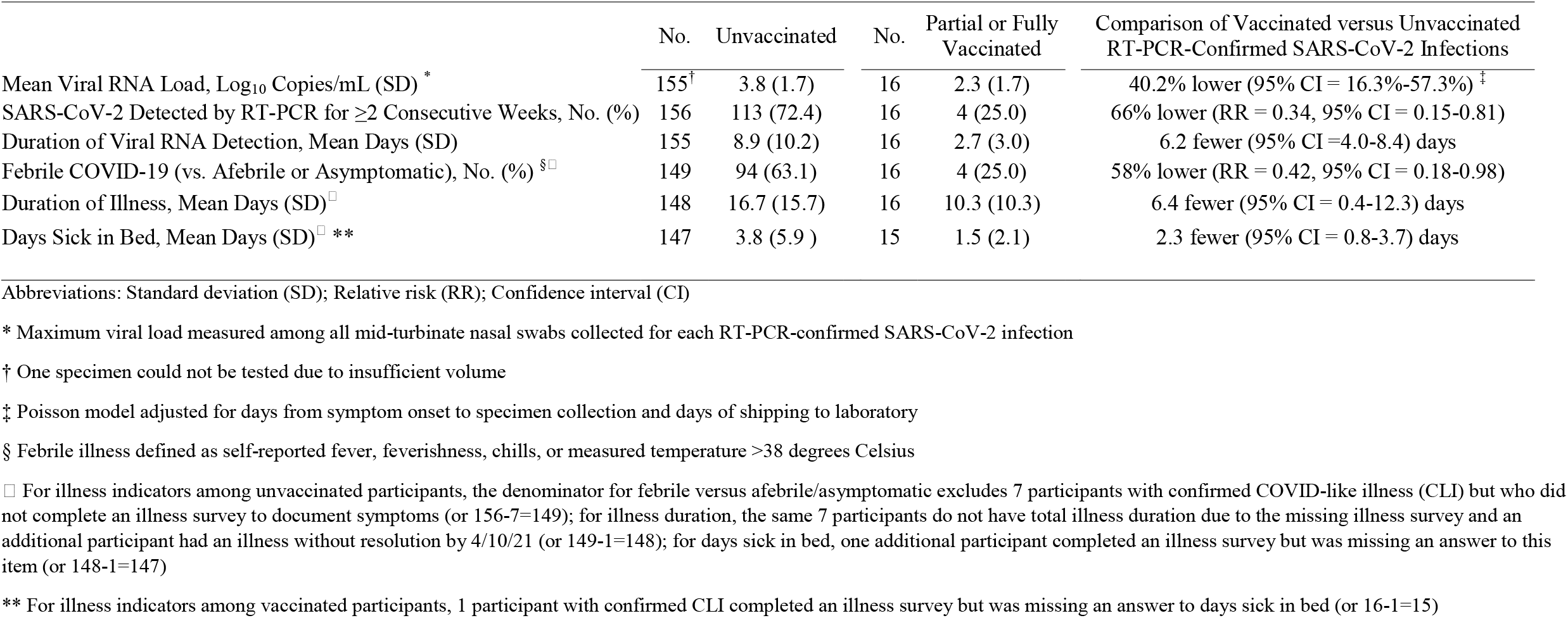
Comparisons between Unvaccinated and Partial or Fully mRNA Vaccine Vaccinated Participants with RT-PCR-Confirmed SARS-CoV-2 Infections in Viral RNA

|  | No. | Unvaccinated | No. | Partial or Fully Vaccinated | Comparison of Vaccinated versus Unvaccinated RT-PCR-Confirmed SARS-CoV-2 Infections |
| --- | --- | --- | --- | --- | --- |
| Mean Viral RNA Load, Log<sub>10</sub> Copies/mL (SD) * | 155<sup>†</sup> | 3.8 (1.7) | 16 | 2.3 (1.7) | 40.2% lower (95% CI = 16.3%-57.3%) <sup>‡</sup> |
| SARS-CoV-2 Detected by RT-PCR for ≥2 Consecutive Weeks, No. (%) | 156 | 113 (72.4) | 16 | 4 (25.0) | 66% lower (RR = 0.34, 95% CI = 0.15-0.81) |
| Duration of Viral RNA Detection, Mean Days (SD) | 155 | 8.9 (10.2) | 16 | 2.7 (3.0) | 6.2 fewer (95% CI =4.0-8.4) days |
| Febrile COVID-19 (vs. Afebrile or Asymptomatic), No. (%) <sup>§□</sup> | 149 | 94 (63.1) | 16 | 4 (25.0) | 58% lower (RR = 0.42, 95% CI = 0.18-0.98) |
| Duration of Illness, Mean Days (SD)<sup>□</sup> | 148 | 16.7 (15.7) | 16 | 10.3 (10.3) | 6.4 fewer (95% CI = 0.4-12.3) days |
| Days Sick in Bed, Mean Days (SD)<sup>□ **</sup> | 147 | 3.8 (5.9 ) | 15 | 1.5 (2.1) | 2.3 fewer (95% CI = 0.8-3.7) days |

Abbreviations: Standard deviation (SD); Relative risk (RR); Confidence interval (CI)
\* Maximum viral load measured among all mid-turbinate nasal swabs collected for each RT-PCR-confirmed SARS-CoV-2 infection
<sup>†</sup> One specimen could not be tested due to insufficient volume
<sup>‡</sup> Poisson model adjusted for days from symptom onset to specimen collection and days of shipping to laboratory
<sup>§</sup> Febrile illness defined as self-reported fever, feverishness, chills, or measured temperature >38 degrees Celsius
<sup>□</sup> For illness indicators among unvaccinated participants, the denominator for febrile versus afebrile/asymptomatic excludes 7 participants with confirmed COVID-like illness (CLI) but who did not complete an illness survey to document symptoms (or 156-7=149); for illness duration, the same 7 participants do not have total illness duration due to the missing illness survey and an additional participant had an illness without resolution by 4/10/21 (or 149-1=148); for days sick in bed, one additional participant completed an illness survey but was missing an answer to this item (or 148-1=147)
\*\* For illness indicators among vaccinated participants, 1 participant with confirmed CLI completed an illness survey but was missing an answer to days sick in bed (or 16-1=15)

The majority of infections among partially or fully vaccinated participants were detected for only a single week (75.0%), while the majority of infections among unvaccinated participants (72.4%) were detected for 2 or more consecutive weeks; this represents a 66% reduction in the relative risk of RT-PCR-detection for ≥2 consecutive weeks (Table 3).

### Vaccine Attenuation of COVID-19–like Illness and Duration

Measures of COVID-19–like illness severity and duration were not associated with participant characteristics, with the exception of a lower mean of symptom duration at Texas and Utah and a lower percentage of febrile COVID-19–like illness at Florida and Utah sites (Table_S6). Among participants with RT-PCR-confirmed SARS-CoV-2 infections, only 25.0% of partially or fully vaccinated participants reported febrile COVID-19–like illness compared to 63.1% of unvaccinated participants; this represents a 58% reduction in the relative risk of febrile COVID-19–like illness following at least partial vaccination (Table 3). Vaccinated participants also reported 6.4 fewer (95% CI =0.4-12.3) total days ill or symptomatic and 2.3 fewer (95% CI=0.8-3.7) days sick in bed with COVID-19–like illness than unvaccinated participants.

## DISCUSSION

In prospective cohorts of 3,975 healthcare personnel, first responders, and other essential and frontline workers followed over 17 weeks in eight U.S. locations, mRNA COVID-19 vaccines (Pfizer-BioNTech’s BNT162b2 and Moderna’s mRNA-1273) were 91% (95% CI = 76%–97%) effective in preventing symptomatic and asymptomatic RT-PCR-confirmed SARS-CoV-2 infection; VE of partial vaccination was 81%. These estimates of VE in real-world conditions are consistent with findings from ^1,2^ and from a similar prospective study of healthcare personnel that also conducts routine SARS-CoV-2 testing ^7^.

Among small number of participants with breakthrough RT-PCR-confirmed SARS-CoV-2 infections despite vaccination, mRNA vaccines appeared to attenuate infection and disease in multiple ways. Specimens from participants who were partially or fully vaccinated at the time of infection had a 40% reduction in viral RNA detected and were less likely to be RT-PCR-positive for more than one week compared to infected, unvaccinated participants. The risk of infected individuals developing febrile COVID-19–like illness was 58% lower and illnesses were on average about 6 days shorter with 2 fewer days sick in bed among partially or fully vaccinated participants compared with unvaccinated participants. Reduced viral RNA presence following mRNA COVID-19 vaccination is consistent with another recent report^8^, and the combination of virologic and clinical effects we observed is consistent with previous findings of lower quantity and duration of viral RNA detection with milder COVID-19 ^9^.

The mechanisms by which COVID-19 vaccination elicits disease attenuation are largely unknown but are likely due to recall of immunologic memory responses that reduce viral replication and accelerate elimination of virally infected cells ^10^. The biologic plausibility of these benefits are supported by similar phenomena observed in other studies ^10,11,12,13,14,15,16,17,18,19^. Our findings are also consistent with reports of lower symptom severity among those with moderate COVID-19 following vaccination with the Ad26.COV2.S COVID-19 vaccine compared to those who received placebo in a randomized controlled trial ^20^.

Among this study’s strengths are the focus on working-age adults without prior laboratory-documented SARS-CoV-2 infections, use of weekly testing for SARS-CoV-2 infection and illness with high surveillance adherence, multi-method documentation of vaccination status, and estimation of VE using vaccination propensity weighting, continuous local virus circulation updates, potential virus exposure, and PPE use. The study’s use of a standard synthetic RNA to conduct quantitative RT-PCR improves upon most prior studies that have relied upon cycle thresholds from real-time RT-PCR as a proxy for viral RNA load ^9^.

This study also has at least seven limitations. First, although our estimate of 81% VE for partial vaccination is similar to other reports ^1,2,7,21,22^, this estimate is based on a relatively brief period of follow-up (median of 19 days partially vaccinated versus 69 days fully vaccinated per participant). Second, we may have overestimated VE if we disproportionately failed to detect infections among vaccinated participants due to vaccine attenuation of viral RNA load or reductions in RT-PCR sensitivity due to self-collection and shipping of specimens ^23^. Third, the study has not completed genetic sequencing for all viruses. Fourth, due to the relatively small number of breakthrough infections observed, we could not differentiate attenuation effects associated with partial versus full vaccination. Similarly, sparse data reduced the precision of estimates, though the consistency of trends across measures affirms the direction of the overall effect. Fifth, due to sparse data and limited racial and ethnic diversity, we were also unable to fully examine or adjust for potential confounders of vaccine attenuation effects. Nonetheless, we stratified our participant recruitment to ensure a combination of participant characteristics by occupation, age, and sex; we did not observe consistent associations between socio-demographic, health, virus exposure, or PPE use with either vaccination status, viral RNA load, or illness duration. Sixth, febrile symptoms and illness duration were measured using self-report, which can be subject to recall and confirmation biases. Yet, these findings are consistent with our virologic findings of reduced viral load and duration of detection. Finally, detection of viral RNA is not equivalent to isolation of infectious virus; however, lower RT-PCR cycle thresholds have been associated with the ability to isolate SARS-CoV-2 in culture^9^, and both viral quantity and duration of viral RNA detection are associated with infectivity and transmission in other viral infections^19,24–26^.

If further data confirms that mRNA vaccination reduces the number of viral RNA particles and the duration of detection and this in turn blunts the infectivity of SARS-CoV-2, then mRNA vaccines are not only highly effective in preventing SARS-CoV-2 infection, but they may also mitigate the impact of breakthrough infections, which is especially important to essential and frontline workers given their potential to transmit the virus through frequent close contacts with patients, co-workers, and the public.

Disclosure forms provided by the authors are available with the full text of this article at NEJM.org.

## Data Availability

Summary data will be available once all study objectives are met.

## ACKNOWLEDGEMENTS

HEROES-RECOVER Network includes Arizona Healthcare, Emergency Response and Other Essential Workers Surveillance Study (HEROES) and Research on the Epidemiology of SARS-CoV-2 in Essential Response Personnel (RECOVER).

Funding provided in whole or in part by federal funds from the National Center for Immunization and Respiratory Diseases, Centers for Disease Control and Prevention under contract numbers 75D30120R68013 awarded to Marshfield Clinic Research Laboratory, 75D30120C08379 to University of Arizona, and 75D30120C08150 awarded to Abt Associates, Inc.

## Disclosures

Allison L. Naleway reported funding from Pfizer for a meningococcal B vaccine study unrelated to the submitted work. Kurt T. Hegmann serves at the Editor of the American College of Occupational and Environmental Medicine’s evidence-based practice guidelines. Matthew S. These reported grants and personal fees from Reed Group and the American College of Occupational and Environmental Medicine, outside the submitted work. Other authors have reported no conflicts of interest

On behalf of HEROES-RECOVER Network, we thank Al Barskey, Lenee Blanton, Christopher Braden, William Brannen, Joseph Bresee, Erin Burns, Joanne Cono, Gordana Derado, Jill Ferdinands, Anthony Fiore, Katherine Garvin, Jacqueline Gindler, Susan Goldstein, Luis Rivera Gonzalez, Brendan Flannery, Aron Hall, Lauri Hicks, Pellumbeshe Hoxhaj, Douglas E. Jordan, Zoe Kaplan, Pam Kennedy, Brian A. King, Archana Kumar, Adam J. Langer, Jennifer Layden, Brandi Limbago, Adam MacNeil, Elissa Meites, Andrea McCollum, L. Clifford McDonald, Christina McMichael, Natalie Olson, Todd Parker, Palak Patel, Mary Reynolds, Sue Reynolds, Stephanie Schrag, Nong Shang, Abigail Shefer, Alan Sims, Robert Slaughter, Dylan Sorensen, Matthew J. Stuckey, Robert V Tauxe, Natalie Thornburg, Vic Veguilla, Jennifer Verani, Katherine (Leza)Young, Rose Wang, Bao-Ping Zhu, CDC; Genesis Barron, Cynthia Beaver, Dimaye Calvo, Esteban Cardona, Adam Carl, Andrea Carmona, Alissa Coleman, Zoe Baccam, Emily Cooksey, Stacy Delgado, Kiara Earley, Ryan Ejindu, Natalie Giroux, Sofia Grijalva, Allan Guidos, Brad Haeckel, Adrianna Hernandez, James Hollister, Theresa Hopkins, Christina Hughey, Rezwana Islam, Gabriella Jimenez, Krystal Jovel, Olivia Kavanagh, Karla Ledezma, Jonathan Leyva, Sally Littau, Amelia Lobos, James Lopez, Paola Louzado Feliciano, Michael Johnson, Elizabeth Kim, Kenneth Komatsu, Veronica Lugo, Jeremy Makar, Taylor Maldonado, Enrique Marquez, Allyson Munoz, Janko Nikolich, Sandrai Norman, Assumpta Nsengiyunva, Kennedy O’brien, Joel Parker, Jonathan Perez Leyva, Alexa Roy, Katerina Santiago, Carlos Silvera, Saskia Smidt, Isabella Terrazas, Tahlia Thompson, Heena Timsina, Erica Vanover, Graham Wegner, Mandie White, April Yingst, University of Arizona, Arizona Department of Health Services, University of Miami; Yolanda Prado, Daniel Sapp, Mi Lee, Chris Eddy, Matt Hornbrook, Danielle Millay, Dorothy Kurdyla, Ambrosia Bass, Kristi Bays, Kimberly Berame, Cathleen Bourdoin, Carlea Buslach, Kenni Graham, Tarika Holness, Abreeanah Magdaleno, Joyce Smith-McGee, Sam Peterson, Aaron Piepert, Krystil Phillips, Joanna Price, Sperry Robinson, Katrina Schell, Emily Schield, Natosha Shirley, Anna Shivinsky, Britta Torgrimson-Ojerio, Shawn Westaway, Kaiser Permanente Northwest; Angela Hunt, Jessica Lundgreen, Karley Respet, Jennifer Viergutz, St. Luke’s; Camie Schaefer, Arlyne Arteaga, Matthew Bruner, Daniel Dawson, Emilee Eden, Jenna Praggastis, Joseph Stanford, Jeanmarie Mayer, Marcus Stucki, Jonathan Thibaudeau, Riley Campbell, Kathy Tran, Madeleine Smith, Braydon Black, Christina Pick, Madison Tallman, Chapman Cox, Derrick Wong, Michael Langston, Adriele Fugal, Fiona Tsang, Maya Wheeler, Gretchen Maughan, Alexis Lowe, University of Utah; Jake Andreae, Adam Bissonnette, Krystal Boese, Michaela Braun, Cody DeHamer, Timothy Dziedzic, Joseph Eddy, Heather Edgren, Wayne Frome, Nolan Herman, Mitchell Hertel, Erin Higdon, Rosebud Johnson, Steve Kaiser, Tammy Koepel, Sarah Kohn, Taylor Kent, Thao Le, Carrie Marcis, Megan Maronde, Isaac McCready, Nidhi Mehta, Daniel Miesbauer, Anne Nikolai, Brooke Olson, Lisa Ott, Cory Pike, Nicole Price, Christopher Reardon, Logan Schafer, Rachel Schoone, Jaclyn Schneider, Tapan Sharma, Melissa Strupp, Janay Walters, Alyssa Weber, Reynor Wilhorn, Ryan Wright, Benjamin Zimmerman, Marshfield Clinic Research Laboratory; Andrea Bronaugh, Tana Brummer, James Carr, Claire Douglas, Michael Duckworth, Kate Durocher, Hala Deeb, Rebecca Devlin, Sauma Doka, Tara Earl, Jini Etolue, Deanna Fleary, Jessica Flores, Chris Flygare, Isaiah Gerber, Louise Hadden, Jenna Harder, Katherine Harris, Don LaLiberty, Lindsay LeClair, Nancy McGarry, Peenaz Mistry, Steve Pickett, Brandon Poe, Khaila Prather, David Pulaski, Rajbansi Raorane, Meghan Shea, Nicole Simmons, Brian Sokol, John Thacker, Matthew Trombley, Pearl Zheng, Chao Zhou, Abt Associates; Alejandro Arroliga, Madhava Beeram, Todd Crumbaker, Thomas Denton, Jason Ettlinger, Angela Kennedy, Mary Kylberg, John Myers, Binitha Rajapudi, Spencer Rose, Natalie Settelie, Rupande Patel, Jennifer Thomas, Baylor Scott & White Health; HEROES-RECOVER cohort participants.

## Footnotes

i Arizona Healthcare, Emergency Response and Other Essential Workers Surveillance Study (HEROES); Research on the Epidemiology of SARS-CoV-2 in Essential Response Personnel (RECOVER).

ii See 45 C.F.R. part 46; 21 C.F.R. part 56

## Notes

### Author Declarations

This study was reviewed and approved by the University of Arizona IRB as the single IRB for this study

### Summary of Updates

Corrected a few critical errors.

