## Supplementary Appendix for "Prevention and Attenuation of COVID-19 by BNT162b2 and mRNA-1273 Vaccines"

### Supplementary Material

**E.** **Laboratory: Genetic sequencing methods………………………………………...14**

**F.** **Statistical analysis methods………………………………………………………...14**

**G.** **Supplemental Tables……………………………………………………………….19**

**H.** **Supplemental Figures……………………………………………………………….33**

**H.** **References…………………………………………………………………………...44**

### Supplementary Appendix Methods

#### A. Protocol Methods for HEROES and applicable to both HEROES and RECOVER cohorts

The HEROES-RECOVER network of cohorts in 8 locations consist of Arizona Healthcare, Emergency Response and Other Essential Workers Surveillance Study (HEROES) and Research on the Epidemiology of SARS-CoV-2 in Essential Response Personnel (RECOVER). An overview of the cohorts' protocol and procedures is available in a pre-publication link for the HEROES platform: **https://pubmed.ncbi.nlm.nih.gov/34057904/**

Although these descriptions are for HEROES, the methods are consistent with RECOVER which shares common protocol and procedures.

##### Participants for HEROES

*Eligibility criteria*

Eligible participants include Arizona residents aged 18–85 years who currently work at least 20 hours per week in an occupation involving regular direct contact (within three feet) with others, assessed at the participant level. We have intentionally chosen a broad occupational category for essential workers in order to capture the full breadth of occupations that cannot socially distance to conduct their work.[1] The occupations are categorized as Health Care Personnel (HCP), First Responders (FR), or Other Essential Workers (OEW). HCP includes clinical providers and support staff in inpatient, outpatient, or residential settings. FR includes firefighters, emergency medical services, law enforcement, border patrol, and correctional officers. OEW includes workers in the following sectors: education, agriculture and food processing, public and other transportation services, solid waste collection, warehouse and delivery, utilities, government and community-based services, childcare, information technology, environmental services, and hospitality. All participants must have access to a smartphone or internet-connected computer, a mailing address, and ability to speak or write English or Spanish. Exclusion criteria include receipt of a COVID-19 vaccine prior to enrollment, although we continue to follow participants who are vaccinated during the study. The majority of the cohort of the healthcare personnel and first responders were recruited prior to vaccine availability.

*Recruitment Strategy*

In order to enroll 4000 participants as quickly as possible, we have employed a multipronged recruitment strategy. First, we are recruiting from ongoing Arizona-based COVID-19 testing activities such as university-driven antibody and saliva testing initiatives and occupation-based state health department surveillance. Second, we have partnered with community-based COVID-19 studies to recruit from ongoing COVID-19 population cohorts. Third, the study accepts self-referrals and so we have developed a marketing strategy to increase general study awareness through press releases and targeted recruitment to occupations.

All recruitment and enrollment activities are conducted remotely utilizing a virtual call-center platform and REDCap[2] to ensure staff and participant safety. Direct recruitment is conducted via phone and email. Participants are given the option to complete a self-screening questionnaire survey that is emailed to them, or to complete a screening interview over the phone. Once deemed eligible and the participant is interested in the study, an electronic consent form is emailed to participants to review and sign electronically through REDCap.

Sampling targets are based upon the employment demographics of Arizona and we seek to enroll essential workers in the following proportions: 50% from 18–40 years old and 50% between 41 and 85 years, 50% women, and 50% Hispanic or American Indian. By occupation, we seek to enroll 40% HCP, 30% FR, and 30% OEW. These sampling breakdowns are presented for every

1000 participants in Table 2. Our goal is to enroll these proportions in both seronegative and seropositive specimens (Table 2). As specified targets are met, recruitment and enrollment priorities will shift to under-enrolled groups.

*Enrollment*

Upon enrollment, participants are asked to complete a baseline questionnaire that collects information about sociodemographic characteristics, health status and behaviors, occupational exposure (tailored to the occupational category), history with and attitudes about COVID-19, and influenza vaccination history during 2020–21 and the previous five seasons (Table 3). Participants are asked to schedule a blood draw (40 mL) within 5 days of enrollment at a laboratory facility in their area in order to complete their baseline serology and are shipped a box of self-collection respiratory supplies so they can begin their active surveillance.

*Active Surveillance*

As part of active surveillance for incident SARS-CoV-2 infection, all participants provide weekly self-collected mid-turbinate nasal swabs appropriate to test for SARS-CoV-2 and influenza (during influenza season). Upon enrollment, study participants are provided information that the study duration could be up to 2 years, but initial expectations are for at least 36 weeks of weekly self-collected respiratory specimens. If an individual experiences COVID-19–like illness (CLI), they are asked to collect an additional respiratory specimen on the date of first CLI symptom onset. Weekly and illness kits are differentiated by color so participants know which to take and study staff can track supplies. Respiratory specimens are analyzed utilizing the CDC-designated reference laboratory for real-time reverse transcription polymerase chain reaction (rRT-PCR) assay testing. AZ HEROES staff will prepare and distribute self-collection kits to the study participants, including detailed paper and video instructions. The laboratory will provide feedback on specimens that were unable to be tested because of participant error in collection or shipping of the sample (e.g. leaking or missing required components). This feedback will be utilized to re-educate participants. If participants receive a positive test result, trained study staff contact participants to provide CDC guidance on quarantine practices and warning signs requiring medical care and answer any questions they may have.

Enrolled participants participate in active surveillance via weekly surveys, explained in detail in the Data Collection section.

#### Data Collection for HEROES

Active surveillance for acute illness is conducted throughout the study period. Participants are prompted to begin surveillance in the week following study enrollment and completion of the baseline survey. Each week, all participants are contacted via text message on their predesignated surveillance day (described in detail below). At the end of each text message exchange, the participant is reminded to collect a weekly specimen on their assigned day for collection.

##### *Active Surveillance surveys*

As a part of active surveillance, participants are contacted weekly via secure short message service (SMS) text messages (via Twilio) asking them two standardized questions about their general health status and presence of CLI symptoms. Twilio is a text-messaging service that can read/write into the study REDCap and customize questions based upon participant responses. In addition to the two standardized questions, each week they receive one of four sets of rotating questions about changes in their occupational SARS-CoV-2 exposure, community and household exposure, and attitudes and beliefs surrounding COVID-19 risk. Any individual who indicates CLI in a weekly survey, or by contacting AZ HEROES staff directly, completes additional information via a mobile-friendly webform including the participant's symptoms, self-reported severity, duration, self-reported medical treatment, during- and post-illness function, and details about the resolution of their illness.

*Self-reported data*

Participants who indicate they have experienced CLI in the last 7 days are moved to an acute illness monitoring flow, where they are instructed to collect and ship an acute illness kit and complete additional questions about their illness episode. Individuals can also be placed into the acute illness monitoring flow by notifying study staff that they are ill. Participants remain in the acute illness arm until they self-report that their illness has resolved. Before returning to the weekly active-surveillance flow, participants complete a recovery survey in which they confirm duration of illness and answer questions about atypical symptoms, productivity loss, and use of health services. Participants continue to take weekly respiratory specimens throughout their acute illness monitoring.

*Vaccine information*

Participants are asked a series of questions to assess their knowledge, attitudes, and practices (KAP) related to SARS-COV-2 vaccination in the enrollment and/or follow-up survey to capture the information prior to vaccination. Similar to previous KAP studies of influenza vaccines,[3,4] participants are asked how much they know about the COVID-19 vaccines, if they received the vaccine, their intention to receive one if they have not, how safe and effective they think the vaccines are, and how likely they are to get sick if they do not receive a vaccine.

As soon as one or multiple COVID-19 vaccines are made available to individuals within the study, they will be prompted about vaccine intent and are asked to text "vaccine" to the text platform when they get vaccinated. Once vaccinated, they complete a brief webform on date of vaccination, vaccine manufacturer and order in sequence (e.g., first or second) for vaccines requiring multiple doses. State Immunization Information System registries will be used as a backup to capture vaccine information about individuals who do not share the information with the study via text message, and for confirmation and completeness on individuals who do receive it.

*Laboratory methods*

**Respiratory specimens.** Participants are asked to self-collect a respiratory specimen each week of the study period. Sampling kits are provided to all study participants, which include collection and shipping supplies for 8 weeks of collections, along with illustrated instructions on how to properly collect and ship their respiratory specimens. Study staff track the use of specimen kits and ship replenishments to participants as needed. Each week, regardless of symptoms, participants collect an anterior mid-turbinate nasal swab on both nares using a flocked swab or equivalent and place it into a tube containing viral transport media (VTM). If participants experience CLI, they use an ‘acute illness kit’ which consists of materials to collect a nasal swab in VTM and a saliva specimen in a saliva-collection tube without stabilixin. All specimens are shipped with a cold pack, using priority overnight express shipping to a CDC-designated laboratory following International Air Transport Association (IATA) guidelines.[5] Upon receipt by the laboratory, specimens are aliquoted and analyzed for SARS-CoV-2 using a rRT-PCR method[6] under FDA emergency use authorization (EUA). Remaining aliquots are maintained for additional analysis, banking or long-term storage.

**Blood specimens.** All participants contribute 40 mL of whole blood at enrollment, at 11- to 13-week intervals, and following a positive rRT-PCR or vaccination events (Figure 1). Participants can submit specimens at participating laboratories closest to the participant’s residence or work. If a participant does not develop symptoms, but SARS-CoV-2 is detected in a weekly specimen, participants are instructed to submit a blood sample approximately 4 weeks following the date of first rRT-PCR detection; if the participant experiences CLI within 2 weeks of virus detection, they are instructed to submit a blood sample 4 weeks after initial symptom onset. If the participant has a convalescent blood specimen drawn prior to another planned repeat blood collection, the scheduling of following blood collections will be 11–13 weeks following the convalescent draw. Participants who receive the COVID-19 vaccine during the study period are asked to provide a blood specimen at 14–21 days after each dose of the vaccine (with the first postvaccination blood draw collected prior to the second vaccination dose, if relevant), and then every 11–13 weeks as described above. Information on adverse events and symptoms related to vaccination will be collected retrospectively after participants receive both doses of the vaccine.

Whole blood is collected and processed using CDC guidelines for serum collection.[7] The serum specimens are divided into aliquots labeled with the same study identification number (Study ID) and specimen ID on all tubes, and an aliquot ID unique to each tube. All specimens are stored at −70 C or colder prior to SARS-CoV-2 antibody analysis or long-term storage. At the University of Arizona, the serum is tested for antibodies against the receptor binding domain (RBD) of the spike protein and verified with the S2 domain of S protein antibodies, as previously described,[8] using the FDA EUA (ID#201116) test. This testing at study entry is used to ensure correct placement of AZ HEROES participants into seronegative or seropositive groups.

*Data collection and security*

Most research activities occur through electronic communications (email, text, and internet-based surveys), telephone contacts, or via postal or express mail, minimizing direct contact between study staff and participants. All surveys are self-administered by participants on a computer or smartphone. Surveys can also be administered by telephone or mail should participants be unable or become unwilling to access them online. Participant information given to study staff via phone or email conversation is entered and stored in REDCap by study staff. Alternatively, data are imported into REDCap from Twilio for participant responses to text surveillance or direct participant response in REDCap.

*Data management*

**REDCap.** A study database is maintained in REDCap. Tracking databases with patient identifiers and contact information are kept securely according to the University of Arizona standard operating procedures with respect to cybersecurity, privacy, patient confidentiality, and compliance with applicable patient privacy regulations. Any study-related documents with personal identifiers are stored in a locked cabinet in lockable offices on campus. All study-related documents and specimens contain a unique identifier for each participant. Data entry forms provide some quality assurance using logic and range checks and automated skip patterns. The research team performs additional data quality checks on a weekly basis, including assessments of missing data. Laboratory results are entered directly into the REDCap study database from the study reference laboratory, including results from rRT-PCR assays and serologic assays. If a reference laboratory is not able to enter data directly, the laboratory is provided a laboratory results reporting template that is then merged with study data using the Specimen ID.

**Twilio.** Twilio is a cloud-based communications platform that allows for automated text messaging chains to be sent to study participants. It is used to send weekly and illness monitoring questions to participants. Participant responses are stored in Twilio until sent as a batch to REDCap once per day.

*Statistical considerations*

**Power Analysis.** Our goal is to recruit 4000 participants, split evenly between seronegative and seropositive individuals. Among the seronegative cohort, we estimated that a sample of >852 is required to achieve 80% power (alpha = .05) to detect a true incidence of SARS-CoV-2 infection of 4% (and the enrolled cohort exceeds this sample estimate at the drafting of this report); thus we expect to be sufficiently powered to make overall estimates and estimates by two-level-strata (such as age, sex, or healthcare personnel vs. others). Power estimation for COVID-19 vaccine effectiveness (VE) was performed using Monte Carlo simulation to generate survival time over 12-months based on varying vaccine coverage (with quarterly increases in 2-dose vaccine coverage from 0% to 80% among HCP, 70% for FR, and 30% for OEW) and varying SARS-CoV-2 incidence rate (from 0.67% to 1.42% monthly attack rate) using the equations proposed by Austin and a Cox marginal model.[9] Based on 1000 simulations, with 2000 participants in the seronegative stratum, the study is estimated to have >80% power to detect a true VE of 75%. If the data are pooled with similar studies using common methodologies to a total of 5000 participants, the combined analysis is estimated to have 99% power to detect a true VE of 75% using the same assumptions.

**Data analysis.** To estimate the incidence of SARS-CoV-2 infection and the corresponding 95% confidence intervals in essential workers, we will fit negative binomial regression models to the data stratified by RT-PCR-confirmed infections, occupation, symptom presentation, close contact exposure, and demographic variables, with follow-up time as an offset. Logistic regression and negative binomial models will be used to estimate the risk of infection in different occupational groups. In the logistic regression model, we will include the log-transformed person weeks as the offset. The model is then adjusted by symptom presentation, demographic factors, study site, and healthcare utilization. The VE (1 − confirmed cases of COVID-19 illness per 1000 person-weeks among vaccinated essential workers ÷ confirmed cases of COVID-19 illness per 1000 person-weeks among unvaccinated essential workers × 100%) with 95% confidence intervals will be estimated by a negative binomial regression model. The potential confounders such as study site and previously seropositive status will be included in the model. We will apply nonlinear mixed models to describe individual and group mean trajectories in neutralizing antibody titers over time. We will classify and identify subgroups of cases by self-reported clinical severity, healthcare utilization, occupational and community exposures, and duration of symptoms. These models will help elucidate the patterns of serologic immunity.

**Ethical considerations**

The study protocol has been reviewed and approved by the University of Arizona Institutional Review Board (IRB). This study was reviewed and approved by the Arizona Department of Health and the University of Arizona's IRBs.[1] CDC and Arizona Department of Health Services (ADHS) IRBs have reviewed the project. The ADHS IRB has approved the project and the CDC IRB deferred to the University of Arizona IRB. The college of public health at the University of Arizona houses all IRB and required study documentation. All participants complete informed consent electronically through the REDCap study database system. Research staff verify participants understand key study activities, are aware of risks, and agree to participate prior to countersigning to confirm consent. Participants receive the results of their weekly and illness COVID-19 tests as well as the results of their antibody testing.

[1] § See 45 C.F.R. part 46.114

#### B. Vaccination Status Documentation

The 796 unvaccinated participants in Table 1 includes 39 participants who received Johnson & Johnson COVID-19 vaccine, but only contribute unvaccinated person-days and are censored from analysis starting on the date of vaccination. Of the remaining 757 unvaccinated participants, 689 (91.0%) participants were confirmed as unvaccinated by multiple methods, including electronic or telephone surveys (at all sites) and reviews of electronic medical and occupational records and/or state immunization registries at sites in Minnesota, Oregon, Texas, and Utah. The remaining 68/757 (9.0%) were at the Arizona or Florida study sites and could not be reached for confirmation. They are presumed to be unvaccinated but are removed from VE estimates in a sensitivity model described below.

#### C. Laboratory Real-time RT-PCR

RNA extraction was performed using the MagMAX Viral/Pathogen Nucleic Acid Isolation Kit on the KingFisher Flex system. RT-PCR was performed using the TaqPath™ COVID-19 Combo Kit on the QuantStudio 7 Pro real time RT-PCR system. Positive specimens were defined as having at least two SARS-CoV-2 targets (ORF1ab, N gene, S gene) with a threshold cycle (Ct) value ≤37 per manufacturer's instructions.[10] Approximately 20% of specimens were randomly selected for retesting as part of routine quality control testing procedures.

#### D. Laboratory: Quantitative SARS-CoV-2 RT-PCR

Residual positive specimens from the Marshfield Clinical Research Institute were frozen at -80 degrees Celsius and shipped on dry ice to the Wisconsin State Laboratory of Hygiene (WSLH) for quantitative SARS-CoV-2 RT-PCR. At the WSLH, specimens were extracted using a QIAcube HT with QIAmp 96 Virus extraction kit (PN 57731) and run on an ABI 7500 Fast Dx using the CDC Influenza SARS-CoV-2 (Flu SC2) Multiplex Assay (https://www.fda.gov/media/139743/download). The SARS-CoV-2 and RNase P targets from this multiplex assay were utilized, and the influenza A and influenza B targets were not analyzed. This assay has Emergency Use Authorization as a qualitative real-time RT-PCR test. To make this assay into a quantitative test, a standard curve of synthetic SARS-CoV-2 RNA (PN 102024, Twist Bioscience) was included on every ABI 7500 run. Starting with 1E+6 copies/µl, a 6-point standard curve of 10-fold dilutions were included on each PCR run, with each dilution run in triplicate (18 wells total). Initially each specimen was also run in triplicate, but because replicates of each specimen were very similar to each other (Ct Standard Deviation <1), after the first run specimens were tested once (in one well). The average Ct values of each dilution of standard were plotted using linear regression, and the linear regression equation was used to convert Ct values of specimens into log copies/µL for each specimen. Specimens with Ct values outside the standard curve were reported as <10 copies/µL or >1,000,000 copies/µL.

For quality control, one negative control and one quantified positive control (Cat. NATSARS COV2-ERC, Zeptometrix Corp.) were included for each extraction run and for each RT-PCR run. For a run to pass, the negative control must be negative, and the positive control must be within the range of mean+/- 3 standard deviations of the average Ct value of the positive control. In addition, to pass quality control the R-squared of the standard curve must be >0.97. In reality, the R-squared was consistently ≥0.99. For each specimen to pass, the RNase P Ct needed to be <35, indicating adequate human specimen collection; all specimens passed this minimal indicator of specimen quality.

Genetic sequence of the SARS-CoV-2 target region were analyzed to determine if genetic substitutions may have impacted genome copy calculations in vaccinated infections. No systematic substitutions were seen in the conserved SC2 target region.

#### E. Laboratory: Genetic sequencing

Available specimens with <32 Ct value by RT-PCR were subjected to SARS-CoV-2 whole genome sequencing by Illumina MiSeq platform following previously published protocols [11]. Additional RT-PCR amplicon amplification followed by Sanger sequencing was applied to the samples with incomplete genome sequences after initial Miseq sequencing.[11] Consensus sequences were generated with Iterative Refinement Meta-Assembler (IRMA) (IRMA v1.0.2 with LABEL v0.6.3 for Linux & Mac OS X, 03-2021) and SARS CoV-2 genome sequence lineage call was based on PANGOLIN v2.3.8 (https://github.com/cov-lineages/pangolin). Lineages were categorized as variants of concern, variants of interest, or wild type or other variants according to criteria published by US Centers for Disease Control and Prevention: SARS-CoV-2 Variant Classifications and Definitions: https://www.cdc.gov/coronavirus/2019-ncov/cases-updates/variant-surveillance/variant-info.html

Sequencing was conducted on SARS-CoV-2 viruses isolated from 22 participants who were ≥7 days post-dose-1 at infection (through March 3 2021) and among 3-4 location- and closest date-matched unvaccinated participants, as available. Due to the very low number of participants with SARS-CoV-2 infection after vaccination and high vaccination uptake among participants, four unvaccinated cases at the same location with infection dates closest to the index case were not always available. A total of 71 unvaccinated participants were identified (**Table S3**).

#### F. Statistical Analysis Methods

Sample Size and Participant Inclusion

As stated in the synopsis from the HEROES protocol, we achieved sufficient sample size >852 seronegative participants required to achieve 80% power (alpha = .05) to detect a true incidence of SARS-CoV-2 infection of 4% (and the enrolled cohort exceeds this sample estimate at the drafting of this report). Of 6,168 eligible participants enrolled, 1,046 withdrew or were lost to follow up prior to December 14, 2020 (**Figure S2**). Withdrawn participants were significantly more likely to be younger, male, not white, Hispanic, and less likely to have a chronic condition (**Table S2**).

Inverse propensity of treatment weighting

Data was divided into weeks and participants were considered immunized if they attained 14 days after vaccination in that week. Baseline covariates in the propensity model included site, sex, age, race, ethnicity, occupation, health status, medical conditions and medications, household characteristics, and influenza vaccination history (**Table S2**).[12] Time varying covariates in the propensity model included study week, local SARS-CoV-2 circulation (percent positive provided by HHS Protect Public Data Hub https://protect-public.hhs.gov), number of hours worked in contact with patients or the public, number of hours in direct contact with someone with known or suspected COVID-19, and percent of time wearing PPE during each of those exposure categories. Local SARS-CoV-2 circulation reflects the average for each week by site. Exposure and PPE use are updated by participants every three weeks if they have changed and the weekly data structure reflects updates as they occur. Propensity to be vaccinated was estimated using boosted regression trees. Average treatment effect (ATE) weights were calculated to assess covariate balance before and after weighting using standardized mean differences. The marginal probability of vaccination was estimated with baseline covariates to stabilize the weights. Final stabilized weights had a mean of 0.95 and maximum of 5.55.

Cox Model

Hazard ratios were calculated by the Andersen-Gill model and vaccine effectiveness was then calculated as 100%×(1-hazard ratio). The Andersen-Gill model is a generalized Cox proportional hazard model that defines the risk intervals based on the counting process. By applying the counting process method, it is possible to model time-to-event data that one can contribute multiple risk intervals [13]. Cox models were weighted using the stabilized weights and had site, local SARS-CoV-2 circulation, and occupation as covariates apriori to adjust for any remaining bias. Robust standard errors were used to account for the clustering by participant created by the stabilized weights.

Assumption of proportional hazard

The proportional hazards assumption was checked for the main and subgroup Cox models by examining correlation between Schoenfeld residuals and time. No evidence of a non-zero slope was found with $p>0.05$ for all tests.

Vaccine attenuation and duration

Attenuation of disease was analyzed among participants with an RT-PCR confirmed infection. We collapsed vaccination exposure into any vaccination due to the small number of breakthrough infections. All analyses compared any vaccination to unvaccinated at the time of illness start. The highest viral RNA loads ($Log_{10}$ copies/mL) measured during RT-PCR-confirmed infections comparisons used a Poisson model. Dichotomous outcomes, PCR positivity for more than one week and febrile illness, used log-logistic regression to calculate relative risks. Comparisons of illness duration outcomes in days were made with Student’s t-test assuming unequal variances. Bivariate analyses assessed baseline characteristics and outcomes for potential relationships and use as covariates. The Poisson model for viral load adjusted for days from symptom onset to specimen collection and for days in transit to the laboratory apriori. Other potential covariates were added independently to each model and kept only if they adjusted the estimate by at least 5%.

Handling of Missing Data

All baseline covariates in the propensity models had complete data. Hours of exposure and percent PPE use are answered by participants when applicable. "Not applicable" is used as a valid response in the boosted regression model and all participant data is used.

VE sensitivity analysis

A sensitivity analysis for VE was conducted censoring person-days associated with potential misclassification bias and with periods of low virus circulation. Specifically, VE was calculated censoring person-time for 68 participants missing confirmation that they were unvaccinated (censoring at the date when the first participant achieved partial immunization at that site location) and 5 participants with an indeterminate RT-PCR result (censoring at the date of symptom onset or collection date for this potentially false negative result). In this model, person-days were also excluded at a study location if local virus circulation fell below 3% for at least 5 days and there were no RT-PCR-confirmed infections in the cohort (**Table S4)**The use of a circulation threshold of 3%-5% has been used in prior VE studies to define seasonal circulation vaccine-preventable viruses in multi-site studies.[14] As listed in the Table below, this applied to 3 study locations; all 3 renewed contribution to person-time at the onset of a new RT-PCR-confirmed SARS-CoV-2 infection among a participant.

| Location | Date reference counties' % positive drops below %3 for ≥5 days | Later date of last study RT-PCR+ and when local positivity drops below 3% [suspension of site person-time] | Date of RT-PCR+ after a site suspension of person-time | Date site person-time starts again after end or break in person-time |
|---|---|---|---|---|
| Phoenix, AZ | Not occurred | No suspension | N/A | N/A |
| Tucson, AZ | 3/28/2021 | 3/28/2021 | 4/5/2021 | 4/5/2021 |
| Other, AZ | Not occurred | No suspension | N/A | N/A |
| Miami, FL | Not occurred | No suspension | N/A | N/A |
| Duluth, MN | 2/15/2021 | 2/15/2021 | 4/5/2021 | 4/5/2021 |
| Portland, OR | 3/6/2021 | 3/6/2021 | 3/30/2021 | 3/30/2021 |
| Temple, TX | Not occurred | No suspension | N/A | N/A |
| Salt Lake City, UT | Not occurred | No suspension | N/A | N/A |

**Table_S1. Comparison of participants included in the vaccine effectiveness analysis population with participants withdrawn or lost to follow up prior to vaccine availability**

| | Total Eligible and Consented Participants N (Col%) | VE Analytic Population N (Row%) | Withdrawn or Lost to Follow-up N (Row%) | p-value* |
|---|---|---|---|---|
| All participants | 5021 (100) | 3975 (79.2) | 1046 (20.8) | |
| Cohort location | | | | <0.0001 |
| Phoenix, AZ | 642 (12.8) | 504 (78.5) | 138 (21.5) | |
| Tucson, AZ | 1561 (31.1) | 1223 (78.3) | 338 (21.7) | |
| Other, AZ | 405 (8.1) | 291 (71.9) | 114 (28.1) | |
| Miami, FL | 355 (7.1) | 239 (67.3) | 116 (32.7) | |
| Duluth, MN | 491 (9.8) | 456 (92.9) | 35 (7.1) | |
| Portland, OR | 528 (10.5) | 491 (93.0) | 37 (7.0) | |
| Temple, TX | 385 (7.7) | 302 (78.4) | 83 (21.6) | |
| Salt Lake City, UT | 654 (13.0) | 469 (71.7) | 185 (28.3) | |
| Sex | | | | <0.0001 |
| Female † | 3041 (60.6) | 2464 (81.0) | 577 (19.0) | |
| Male | 1980 (39.4) | 1511 (76.3) | 469 (23.7) | |
| Age (Years) | | | | <0.0001 |
| 18-49 | 3724 (74.2) | 2847 (76.5) | 877 (23.5) | |
| ≥50 | 1297 (25.8) | 1128 (87.0) | 169 (13.0) | |
| Race | | | | <0.0001 |
| White | 4226 (84.2) | 3431 (81.2) | 795 (18.8) | |
| Other | 795 (15.8) | 544 (68.4) | 251 (31.6) | |
| Ethnicity | | | | <0.0001 |
| Hispanic/Latinx | 992 (19.8) | 685 (69.1) | 307 (30.9) | |
| Other | 4029 (80.2) | 3290 (81.7) | 739 (18.3) | |
| Occupation ‡ | | | | <0.0001 |
| Primary HCP | 919 (18.3) | 809 (88.0) | 110 (12.0) | |
| Nurses and other allied HCP | 1619 (32.2) | 1310 (80.9) | 309 (19.1) | |
| First Responders | 1161 (23.1) | 818 (70.5) | 343 (29.5) | |
| Essential and other frontline | 1271 (25.3) | 1038 (81.7) | 233 (18.3) | |
| Missing | 51 (1.0) | 0 (0.0) | 51 (100.0) | |
| Chronic Condition | | | | <0.0001 |
| None§ | 3623 (72.2) | 2728 (75.3) | 895 (24.7) | |
| 1 or more | 1398 (27.8) | 1247 (89.2) | 151 (10.8) | |

Abbreviations: Vaccine effectiveness (VE), Healthcare personnel (HCP)

*P-values calculated for categorical variables using Pearson's chi-squared test or Fisher's exact test for cells with <5 observations.

† For 58 participants missing biological sex, it was imputed as the more common category, female.

‡ Occupation categories: Primary HCP (physicians, physician assistants, nurse practitioners, dentists), Other allied HCP (nurses, therapists, technicians, medical assistants, orderlies and all others providing clinical support in inpatient or outpatient settings), first responders (FR; firefighters, law enforcement, corrections, emergency medical technicians), essential and frontline workers (EFW; workers in hospitality, delivery, and retail; teachers; all other occupations that require contact within 3 feet of the public, customers, or co-workers as a routine part of their job).

§ For 297 participants, who did not respond to the self-reported question, they were imputed as none, pending further verification.

**Table_S2. Extended version of Table 1 with characteristics of participants by percentage with RT-PCR-confirmed SARS-CoV-2 infections and percentage receiving ≥1 dose of messenger RNA COVID vaccine during study period; all variables contributed to immunization propensity weight calculations**

| | Unique Participants | SARS-CoV-2 PCR-Negatives | SARS-CoV-2 PCR-Positives | | Unvaccinated | Vaccinated with ≥1 Dose | |
|---|---|---|---|---|---|---|---|
| | N ( Col. % ) | N ( Row % ) | N ( Row % ) | P-value | N ( Row % ) | N ( Row % ) | P-value |
| All participants † | 3975 | 3771 ( 94.9 ) | 204 ( 5.1 ) | | 796 ( 20.0 ) | 3179 ( 80.0 ) | |
| Socio-demographic characteristics | | | | | | | |
| Cohort location[‡,§] | | | | <0.0001 | | | <0.0001 |
| Phoenix, AZ | 504 ( 12.7 ) | 461 ( 91.5 ) | 43 ( 8.5 ) ‡ | | 105 ( 20.8 ) | 399 ( 79.2 ) | |
| Tucson, AZ | 1223 ( 30.8 ) | 1148 ( 93.9 ) | 75 ( 6.1 ) ‡ | | 274 ( 22.4 ) | 949 ( 77.6 ) | |
| Other, AZ | 291 ( 7.3 ) | 276 ( 94.8 ) | 15 ( 5.2 ) ‡ | | 70 ( 24.1 ) | 221 ( 75.9 ) | |
| Miami, FL | 239 ( 6.0 ) | 216 ( 90.4 ) | 23 ( 9.6 ) ‡ | | 111 ( 46.4 ) | 128 ( 53.6 ) | |
| Duluth, MN | 456 ( 11.5 ) | 445 ( 97.6 ) | 11 ( 2.4 ) | | 32 ( 7.0 ) | 424 ( 93.0 ) | |
| Portland, OR | 491 ( 12.4 ) | 486 ( 99.0 ) | 5 ( 1.0 ) | | 44 ( 9.0 ) | 447 ( 91.0 ) | |
| Temple, TX | 302 ( 7.6 ) | 284 ( 94.0 ) | 18 ( 6.0 ) ‡ | | 66 ( 21.9 ) | 236 ( 78.1 ) | |
| Salt Lake City, UT | 469 ( 11.8 ) | 455 ( 97.0 ) | 14 ( 3.0 ) | | 94 ( 20.0 ) | 375 ( 80.0 ) | |
| Sex | | | | 0.0240 | | | <0.0001 |
| Female[ǀ] | 2464 ( 62.0 ) | 2349 ( 95.5 ) | 111 ( 4.5 ) | | 423 ( 17.2 ) | 2037 ( 82.8 ) | |
| Male | 1511 ( 38.0 ) | 1422 ( 93.9 ) | 93 ( 6.1 ) | | 373 ( 24.6 ) | 1142 ( 75.4 ) | |
| Age (Years) | | | | 0.5122 | | | 0.0051 |
| 18-49 | 2847 ( 71.6 ) | 2705 ( 95.0 ) | 142 ( 5.0 ) | | 602 ( 21.1 ) | 2245 ( 78.9 ) | |
| ≥50 | 1128 ( 28.4 ) | 1066 ( 94.5 ) | 62 ( 5.5 ) | | 194 ( 17.2 ) | 934 ( 82.8 ) | |
| Race | | | | 0.6882 | | | 0.0012 |
| White | 3431 ( 86.3 ) | 3253 ( 94.8 ) | 178 ( 5.2 ) | | 659 ( 19.2 ) | 2772 ( 80.8 ) | |
| Other | 544 ( 13.7 ) | 518 ( 95.2 ) | 26 ( 4.8 ) | | 137 ( 25.2 ) | 407 ( 74.8 ) | |
| Ethnicity | | | | <0.0001 | | | <0.0001 |
| Hispanic/Latinx | 685 ( 17.2 ) | 625 ( 91.2 ) | 60 ( 8.8 ) | | 198 ( 28.9 ) | 487 ( 71.1 ) | |
| Other | 3290 ( 82.8 ) | 3146 ( 95.6 ) | 144 ( 4.4 ) | | 598 ( 18.2 ) | 2692 ( 81.8 ) | |
| Marital status | | | | 0.1368 | | | <0.0001 |
| Married | 2514 ( 63.2 ) | 2375 ( 94.5 ) | 139 ( 5.5 ) | | 437 ( 17.4 ) | 2077 ( 82.6 ) | |

| | | | | | | | |
|---|---|---|---|---|---|---|---|
| Other | 1461 (36.8) | 1396 (95.6) | 65 (4.4) | | 359 (24.6) | 1102 (75.4) | |
| <u>Occupation</u> | | | | | | | |
| Occupation ¶ | | | | <0.0001 | | | <0.0001 |
| Primary HCP | 809 (20.4) | 793 (98.0) | 16 (2.0) | | 45 (5.6) | 764 (94.4) | |
| Nurses and other allied HCP | 1310 (33.0) | 1244 (95.0) | 66 (5.0) | | 204 (15.6) | 1106 (84.4) | |
| First Responders | 818 (20.6) | 745 (91.1) | 73 (8.9) | | 257 (31.4) | 561 (68.6) | |
| Essential and other frontline | 1038 (26.1) | 989 (95.3) | 49 (4.7) | | 290 (27.9) | 748 (72.1) | |
| <u>Household characteristics</u> | | | | | | | |
| Number of bedrooms | | | | 0.6762 | | | 0.0689 |
| 1 | 222 (5.6) | 210 (94.6) | 12 (5.4) | | 55 (24.8) | 167 (75.2) | |
| 2 | 568 (14.3) | 545 (96.0) | 23 (4.0) | | 109 (19.2) | 459 (80.8) | |
| 3 | 1601 (40.3) | 1517 (94.8) | 84 (5.2) | | 322 (20.1) | 1279 (79.9) | |
| 4 | 1473 (37.1) | 1395 (94.7) | 78 (5.3) | | 262 (17.8) | 1211 (86.8) | |
| Unknown/refused | 111 (2.8) | 104 (93.7) | 7 (6.3) | | 48 (43.2) | 63 (60.6) | |
| Other individuals in household | | | | 0.0766 | | | 0.0002 |
| 0 | 517 (13.0) | 483 (93.4) | 34 (6.6) | | 132 (25.5) | 385 (74.5) | |
| 1 | 1016 (25.6) | 968 (95.3) | 48 (4.7) | | 179 (17.6) | 837 (82.4) | |
| 2 | 882 (22.2) | 850 (96.4) | 32 (3.6) | | 174 (19.7) | 708 (80.3) | |
| 3 | 884 (22.2) | 830 (93.9) | 54 (6.1) | | 153 (17.3) | 731 (82.7) | |
| 4 or more | 676 (17.0) | 640 (94.7) | 36 (5.3) | | 158 (23.4) | 518 (76.6) | |
| Children in household | | | | 0.5846 | | | 0.9193 |
| None | 2081 (52.4) | 1978 (95.1) | 103 (4.9) | | 418 (20.1) | 1663 (79.9) | |
| 1 or more | 1894 (47.6) | 1793 (94.7) | 101 (5.3) | | 378 (20.0) | 1516 (80.0) | |
| <u>Health status</u> | | | | | | | |
| Self-Rated Health | | | | 0.0955 | | | <0.0001 |
| Excellent | 966 (24.3) | 905 (93.7) | 61 (6.3) | | 165 (17.1) | 801 (82.9) | |
| Very good | 1810 (45.5) | 1730 (95.6) | 80 (4.4) | | 321 (17.7) | 1489 (82.3) | |
| Good/Fair/Poor | 1199 (30.2) | 1136 (94.7) | 63 (5.3) | | 310 (25.9) | 889 (74.1) | |
| Chronic Condition | | | | 0.8765 | | | 0.0023 |
| None** | 2728 (68.6) | 2589 (94.9) | 139 (5.1) | | 582 (21.3) | 2146 (78.7) | |

| | | | | | | | |
|---|---|---|---|---|---|---|---|
| 1 or more | 1247 (31.4) | 1182 (94.8) | 65 (5.2) | | 214 (17.2) | 1033 (82.8) | |
| Daily medications | | | | 0.5732 | | | <0.0001 |
| 0 | 1931 (48.6) | 1839 (95.2) | 92 (4.8) | | 454 (23.5) | 1477 (76.5) | |
| 1 | 852 (21.4) | 805 (94.5) | 47 (5.5) | | 149 (17.5) | 703 (82.5) | |
| 2 | 533 (13.4) | 499 (93.6) | 34 (6.4) | | 90 (16.9) | 443 (83.1) | |
| 3 | 322 (8.1) | 308 (95.7) | 14 (4.3) | | 41 (12.7) | 281 (87.3) | |
| 4 or more | 337 (8.5) | 320 (95.0) | 17 (5.0) | | 62 (18.4) | 275 (81.6) | |
| Health behaviors | | | | | | | |
| Smoking | | | | 0.9940 | | | 0.0035 |
| Not current smoker | 3099 (78.0) | 2940 (94.9) | 159 (5.1) | | 590 (19.0) | 2509 (81.0) | |
| Smoke tobacco products | 876 (22.0) | 831 (94.9) | 45 (5.1) | | 206 (23.5) | 670 (76.5) | |
| Influenza vaccination history in past 5 years | | | | <0.0001 | | | <0.0001 |
| No vaccination history | 646 (16.3) | 591 (91.5) | 55 (8.5) | | 297 (46.0) | 349 (54.0) | |
| 1 - 3 years of vaccination | 628 (15.8) | 589 (93.8) | 39 (6.2) | | 194 (30.9) | 434 (69.1) | |
| 4 or more years of vaccination | 2701 (67.9) | 2591 (95.9) | 110 (4.1) | | 305 (11.3) | 2396 (88.7) | |
| Potential virus exposures and use of PPE, Median (IQR) of Average Monthly Updates per Participant[††] | | | | | | | |
| Hours within 3 feet of others at work | 27 (20.0-35.3) | 27 (20.0-35.2) | 25 (20.0-37.9) | 0.1031 | 26 (20.0-35.6) | 27 (20.0-35.2) | 0.1056 |
| While in close contact at work, percent time using PPE[‡‡] | 99 (90.0-100) | 99 (90.0-100) | 100 (89.0-100) | 0.6347 | 96 (78.6-100) | 99 (99.4-100) | <0.0001 |
| Hours within 3 feet of suspected or confirmed COVID-19 at work, home, or community | 8 (2.2-24.0) | 8 (2.2-24.0) | 6 (2.0-23.2) | 0.4463 | 10 (3.1-26.7) | 7 (2.0-23.4) | 0.0003 |

Abbreviations: Interquartile range (IQR), Healthcare personnel (HCP), First responders (FR), Messenger RNA (mRNA), Personal protective equipment (PPE)

*P-values calculated for categorical variables using Pearson's chi-squared test or Fisher's exact test for cells with <5 observations; Kruskal-Wallis non-parametric tests was used to compare median values.

† Analytic sample excludes 1,147 participants with documented SARS-CoV-2 infection before enrollment or as part of surveillance.

‡ Sites identified had higher percentages of their participants with RT-PCR-confirmed SARS-CoV-2 infections than the other sites Chi-square = 41.0, p-value <0.0001.

§ Comparison of those who were vaccinated with at least one dose and those who were not, cohort locations for Portland, OR, Duluth, MN, Salt Lake City UT were combined compared to Phoenix, AZ, Tucson, AZ, Other, AZ, Miami, FL and Temple, TX with chi-square value of 88.3 (p-value <0.0001).

‖ For 15 participants missing biological sex, it was imputed as the more common category (female).

¶ Occupation categories: Primary HCP (physicians, physician assistants, nurse practitioners, dentists), Other allied HCP (nurses, therapists, technicians, medical assistants, orderlies and all others providing clinical support in inpatient or outpatient settings), first responders (FR; firefighters, law enforcement, corrections, emergency medical technicians), essential and frontline workers (EFW; workers in hospitality, delivery, and retail; teachers; all other occupations that require contact within 3 feet of the public, customers, or co-workers as a routine part of their job).

** For 77 participants, who did not respond to the self-report question, they were imputed as none, pending further verification.

†† Each month, participants were asked about close contacts and PPE use during the past 7 days. The mean of monthly responses during the study period were calculated.

‡‡ Only applicable for participants indicating a potential exposure during the past 7 days.

**Table_S3. Number and percentage of SARS-CoV-2 viruses by three lineage classifications and by vaccination status at infection and cohort location**

Whole genome sequencing was conducted at CDC using previously published protocols for SARS-CoV-2 viruses detected among 22 participants who were ≥7 days post-dose-1 at infection (through March 3 2021) and among 3-4 unvaccinated participants at the same location with infection dates closest to the index case. Lineages were categorized as variants of concern, interest, or other by CDC website (Supplementary_Appendix_Methods).

| | Variants of Concern | | Variants of Interest | | Wild Type & Other | | |
|---|---|---|---|---|---|---|---|
| | N | ( Col. % ) | N | ( Col. % ) | N | ( Col. % ) | |
| **Total** | **10** | | **1** | | **82** | | |
| By vaccination status at infection | | | | | | | Variants of concern / All (but not variant of interest) |
| Unvaccinated | 7 | ( 70 ) | 1 | ( 100 ) | 63 | ( 77 ) | 7/70 (10%) |
| Indeterminate (days 1-13 post dose-1) | 0 | ( 0 ) | 0 | ( 0 ) | 12 | ( 15 ) | 0/12 (0%) |
| Partially or fully vaccinated (≥14-days post dose-1)* | 3 | ( 30 ) | 0 | ( 0 ) | 7 | ( 9 ) | 3/10 (30%) |
| By cohort location | | | | | | | |
| Phoenix, AZ | 2 | ( 20 ) | 1 | ( 100 ) | 12 | ( 15 ) | |
| Tucson, AZ | 2 | ( 20 ) | 0 | ( 0 ) | 22 | ( 32 ) | |
| Other, AZ | 1 | ( 10 ) | 0 | ( 0 ) | 14 | ( 17 ) | |
| Miami, FL | 0 | ( 0 ) | 0 | ( 0 ) | 5 | ( 1 ) | |
| Duluth, MN | 0 | ( 0 ) | 0 | ( 0 ) | 7 | ( 9 ) | |
| Portland, OR | 0 | ( 0 ) | 0 | ( 0 ) | 2 | ( 2 ) | |
| Temple, TX | 1 | ( 10 ) | 0 | ( 0 ) | 10 | ( 12 ) | |
| Salt Lake City, UT | 4 | ( 40 ) | 0 | ( 0 ) | 10 | ( 12 ) | |
| Month of detection | | | | | | | |

| | | | | | | | | | | | | |
|---|---|---|---|---|---|---|---|---|---|---|---|---|
| December | 1 | ( | 10 | ) | 0 | ( | 0 | ) | 34 | ( | 41 | ) |
| January | 5 | ( | 50 | ) | 1 | ( | 100 | ) | 38 | ( | 46 | ) |
| February | 3 | ( | 30 | ) | 0 | ( | 0 | ) | 10 | ( | 12 | ) |
| March | 1 | ( | 10 | ) | 0 | ( | 0 | ) | 0 | ( | 0 | ) |
| April | 0 | ( | 0 | ) | 0 | ( | 0 | ) | 0 | ( | 0 | ) |
| By cohort location and unvaccinated vs. vaccinated (excluding 12 indeterminates; all wild type or other) | | | | | | | | | | | | |
| Phoenix, AZ | | | | | | | | | | | | |
| Unvaccinated | 2 | ( | 20 | ) | 1 | ( | 100 | ) | 9 | ( | 11 | ) |
| Partially or fully vaccinated | 0 | ( | 0 | ) | 0 | ( | 0 | ) | 2 | ( | 2 | ) |
| Tucson, AZ | | | | | | | | | | | | |
| Unvaccinated | 1 | ( | 10 | ) | 0 | ( | 0 | ) | 18 | ( | 27 | ) |
| Partially or fully vaccinated | 1 | ( | 10 | ) | 0 | ( | 0 | ) | 1 | ( | 1 | ) |
| Other, AZ | | | | | | | | | | | | |
| Unvaccinated | 1 | ( | 10 | ) | 0 | ( | 0 | ) | 10 | ( | 12 | ) |
| Partially or fully vaccinated | 0 | ( | 0 | ) | 0 | ( | 0 | ) | 2 | ( | 2 | ) |
| Miami, FL | | | | | | | | | | | | |
| Unvaccinated | 0 | ( | 0 | ) | 0 | ( | 0 | ) | 4 | ( | 0 | ) |
| Partially or fully vaccinated | 0 | ( | 0 | ) | 0 | ( | 0 | ) | 0 | ( | 0 | ) |
| Duluth, MN | | | | | | | | | | | | |
| Unvaccinated | 0 | ( | 0 | ) | 0 | ( | 0 | ) | 4 | ( | 5 | ) |
| Partially or fully vaccinated | 0 | ( | 0 | ) | 0 | ( | 0 | ) | 2 | ( | 2 | ) |
| Portland, OR | | | | | | | | | | | | |
| Unvaccinated | 0 | ( | 0 | ) | 0 | ( | 0 | ) | 1 | ( | 1 | ) |
| Partially or fully vaccinated | 0 | ( | 0 | ) | 0 | ( | 0 | ) | 0 | ( | 0 | ) |
| Temple, TX | | | | | | | | | | | | |
| Unvaccinated | 1 | ( | 10 | ) | 0 | ( | 0 | ) | 7 | ( | 9 | ) |
| Partially or fully vaccinated | 0 | ( | 0 | ) | 0 | ( | 0 | ) | 0 | ( | 0 | ) |

| | | | | |
|---|---|---|---|---|---|---|---|---|---|---|---|---|
| Salt Lake City, UT | | | | | | | | | | | | |
| Unvaccinated | 2 | ( | 20 | ) | 0 | ( | 0 | ) | 10 | ( | 12 | ) |
| Partially or fully vaccinated | 2 | ( | 20 | ) | 0 | ( | 0 | ) | 0 | ( | 0 | ) |
| By lineage classification from sequencing | | | | | | | | | | | | |
| B.1.429 | 8 | ( | 80 | ) | | | | | | | | |
| B.1.1.7 | 1 | ( | 10 | ) | | | | | | | | |
| B.1.427 | 1 | ( | 10 | ) | | | | | | | | |
| P.2 | | | | | 1 | ( | 100 | ) | | | | |
| B.1 | | | | | | | | | 9 | ( | 11 | ) |
| B.1.1.231 | | | | | | | | | 1 | ( | 1 | ) |
| B.1.1.316 | | | | | | | | | 4 | ( | 5 | ) |
| B.1.1.434 | | | | | | | | | 1 | ( | 1 | ) |
| B.1.2 | | | | | | | | | 42 | ( | 51 | ) |
| B.1.234 | | | | | | | | | 1 | ( | 1 | ) |
| B.1.239 | | | | | | | | | 2 | ( | 2 | ) |
| B.1.243 | | | | | | | | | 6 | ( | 7 | ) |
| B.1.400 | | | | | | | | | 2 | ( | 2 | ) |
| B.1.409 | | | | | | | | | 1 | ( | 1 | ) |
| B.1.517 | | | | | | | | | 1 | ( | 1 | ) |
| B.1.551 | | | | | | | | | 5 | ( | 6 | ) |
| B.1.565 | | | | | | | | | 1 | ( | 1 | ) |
| B.1.587 | | | | | | | | | 1 | ( | 1 | ) |
| B.1.596 | | | | | | | | | 4 | ( | 5 | ) |
| B.1.609 | | | | | | | | | 1 | ( | 1 | ) |

*Among variants of concern, 1 was partially vaccinated, 2 were fully vaccinated. Among wild type and other variants, 6 were partially vaccinated and 1 was fully vaccinated

**Table S4. Sensitivity analysis to main vaccine effectiveness (VE) estimates that eliminates person-time for those with potential vaccination or infection misclassification and during periods of low local virus circulation**

| | Contributing Participants * | Total Person-Days | Median (IQR) Days | SARS-CoV-2 Infections | Unadjusted VE | | Adjusted VE † | |
|---|---|---|---|---|---|---|---|---|
| mRNA COVID-19 vaccination status | | | | | % | ( 95% CI ) | % | ( 95% CI ) |
| Unvaccinated | 3,948 | 121,992 | 17 (8 - 40) | 151 | | | | |
| Partially vaccinated (≥14-days post dose-1 to day 13 post dose-2) | 2,995 | 80,638 | 22 (21 - 28) | 11 | 87 | ( 74 - 93 ) | 81 | ( 64 - 90 ) |
| Fully vaccinated (≥14-days post dose-2) | 2,508 | 159,898 | 69 (52 - 81) | 5 | 92 | ( 80 - 97 ) | 91 | ( 77 - 97 ) |

Abbreviations: Messenger RNA (mRNA), Vaccine effectiveness (VE), Interquartile range (IQR)

* Contributing participants in vaccination categories do not equal the number with each vaccination dose because participants must have met the vaccination criteria for each status category

† Adjusted VE is inversely weighted for propensity to be vaccinated with doubly robust adjustment for local virus circulation, study location, and occupation. This model excludes person-time among those presumed to be unvaccinated but lacking confirmation (n = 68), person-time after an indeterminate RT-PCR result (n = 5), and person-time during weeks of low local virus circulation (defined as no RT-PCR-confirmed infections within local cohort and percent positive of local SARS-CoV-2 testing fell below 3% for ≥5 days): Tucson, AZ suspended 3/28 to 4/5/21; Duluth, MN suspended 2/15 to 4/4/21; Portland, OR suspended 3/6 to 3/31/21. Also see Figure_S1.

**Table S5. Participant characteristics by mRNA vaccine vaccination status at time of RT-PCR-confirmed SARS-CoV-2 infections**

| | All SARS-CoV-2 RT-PCR-Positives | | SARS-CoV-2 Positives by Vaccination Status at Infection | | | | | | | | Partial and Full Vaccination Combined | | | | | |
|---|---|---|---|---|---|---|---|---|---|---|---|---|---|---|---|---|
| | | | Unvaccinated | | Partially Vaccinated | | Fully Vaccinated | | | | Unvaccinated | | Any Vaccination | | | |
| | N | Col. % | N | Row % | N | Row % | N | Row % | | p-value* | N | Col. % | N | Col % | | p-value* |
| All participants [†] | 204 | | 156 | 76.5 | 11 | 5.4 | 5 | 2.45 | | | 156 | 76.5 | 16 | 7.8 | | |
| Socio-demographic characteristics | | | | | | | | | | | | | | | | |
| Cohort location[‡,§] | | | | | | | | | | 0.0031 | | | | | | 0.0182 |
| Phoenix, AZ | 43 | 8.5 | 32 | 74.4 | 3 | 7.0 | 0 | 0.0 | ‡ | | 32 | 20.5 | 3 | 18.8 | § | |
| Tucson, AZ | 75 | 6.2 | 63 | 84.0 | 1 | 1.3 | 2 | 2.7 | ‡ | | 63 | 40.4 | 3 | 18.8 | § | |
| Other, AZ | 15 | 5.2 | 9 | 60.0 | 1 | 6.7 | 1 | 6.7 | ‡ | | 9 | 5.8 | 2 | 12.5 | § | |
| Miami, FL | 23 | 9.7 | 22 | 95.7 | 0 | 0.0 | 0 | 0.0 | ‡ | | 22 | 14.1 | 0 | 0.0 | § | |
| Duluth, MN | 11 | 2.2 | 6 | 54.5 | 3 | 27.3 | 0 | 0.0 | | | 6 | 3.8 | 3 | 18.8 | | |
| Portland, OR | 5 | 0.8 | 2 | 40.0 | 0 | 0.0 | 1 | 20.0 | | | 2 | 1.3 | 1 | 6.3 | | |
| Temple, TX | 18 | 5.7 | 13 | 72.2 | 1 | 5.6 | 0 | 0.0 | ‡ | | 13 | 8.3 | 1 | 6.3 | § | |
| Salt Lake City, UT | 14 | 3.0 | 9 | 64.3 | 2 | 14.3 | 1 | 7.1 | | | 9 | 5.8 | 3 | 18.8 | | |
| Sex | | | | | | | | | | 0.1713 | | | | | | 0.063 |
| Female | 111 | 4.4 | 79 | 71.2 | 8 | 7.2 | 4 | 3.6 | | | 79 | 50.6 | 12 | 75.0 | | |
| Male | 93 | 6.3 | 77 | 82.8 | 3 | 3.2 | 1 | 1.1 | | | 77 | 49.4 | 4 | 25.0 | | |
| Age (Years) | | | | | | | | | | 0.8332 | | | | | | 0.5969 |
| 18-49 | 142 | 4.9 | 107 | 75.4 | 8 | 5.6 | 4 | 2.8 | | | 107 | 68.6 | 12 | 75.0 | | |
| ≥50 | 62 | 5.5 | 49 | 79.0 | 3 | 4.8 | 1 | 1.6 | | | 49 | 31.4 | 4 | 25.0 | | |
| Race | | | | | | | | | | 0.6995 | | | | | | 0.4014 |
| White | 178 | 5.1 | 138 | 77.5 | 9 | 5.1 | 4 | 2.2 | | | 138 | 88.5 | 13 | 81.3 | | |
| Other | 26 | 5.1 | 18 | 69.2 | 2 | 7.7 | 1 | 3.8 | | | 18 | 11.5 | 3 | 18.8 | | |
| Ethnicity | | | | | | | | | | 0.1861 | | | | | | 0.1216 |
| Hispanic/Latinx | 60 | 8.5 | 40 | 66.7 | 4 | 6.7 | 3 | 5.0 | | | 40 | 25.6 | 7 | 43.8 | | |

| | | | | | | | | | | | | | | |
|---|---|---|---|---|---|---|---|---|---|---|---|---|---|---|
| Other | 144 | (4.4) | 116 | (80.6) | 7 | (4.9) | 2 | (1.4) | | 116 | (74.4) | 9 | (56.3) | |
| Occupation[ǀ] | | | | | | | | | 0.0257 | | | | | 0.0278 |
| Primary HCP | 16 | (2.0) | 8 | (50.0) | 3 | (18.8) | 0 | (0.0) | | 8 | (5.1) | 3 | (18.8) | |
| Nurses and other allied HCP | 66 | (4.9) | 45 | (68.2) | 6 | (9.1) | 2 | (3.0) | | 45 | (28.8) | 8 | (50.0) | |
| First Responders | 73 | (9.2) | 62 | (84.9) | 1 | (1.4) | 2 | (2.7) | | 62 | (39.7) | 3 | (18.8) | |
| Essential and other frontline | 49 | (4.5) | 41 | (83.7) | 1 | (2.0) | 1 | (2.0) | | 41 | (26.3) | 2 | (12.5) | |
| Chronic Condition | | | | | | | | | 0.5743 | | | | | 0.7371 |
| None [¶] | 139 | (5.1) | 104 | (74.8) | 6 | (4.3) | 4 | (2.9) | | 104 | (66.7) | 10 | (62.5) | |
| 1 or more | 65 | (5.1) | 52 | (80.0) | 5 | (7.7) | 1 | (1.5) | | 52 | (33.3) | 6 | (37.5) | |
| Potential exposures to virus from monthly reports, Median (IQR)** | | | | | | | | | 0.3571 | | | | | 0.1723 |
| Average hours worked in direct contact with coworkers | 25 | (20.0-37.9) | 25 | (20.0-38.9) | 20 | (20.0-26.2) | 28 | (20.0-28.4) | | 25 | (20.0-38.9) | 20 | (20.0-28.4) | |
| | | | | | | | | | 0.5449 | | | | | 0.6364 |
| Average hours of direct contact with suspected or confirmed SARS-CoV-2 infection | 6 | (2.0-23.2) | 8 | (2.0-20.0) | 3 | (2.2-3.6) | 18.8 | (2.6-30.4) | | 8 | (2.0-20.0) | 3 | (2.2-30.0) | |
| Use of personal protective equipment (PPE) from monthly reports | | | | | | | | | | | | | | |
| PPE use during work [††] | | | | | | | | | N/A | | | | | N/A |
| No | 0 | (0.0) | 0 | (0.0) | 0 | (0.0) | 0 | (0.0) | | 0 | (0.0) | 0 | (0.0) | |
| Yes | 163 | (79.9) | 123 | (75.5) | 10 | (6.1) | 5 | (3.1) | | 123 | (78.8) | 15 | (93.8) | |
| Missing | 41 | | 33 | | 1 | | 0 | | | | | | | |
| PPE use at work, community, home [††] | | | | | | | | | 0.0532 | | | | | 0.0202 |
| No close SARS-CoV-2 contact in past 7 days | 84 | (41.2) | 66 | (78.6) | 2 | (2.4) | 0 | (0.0) | | 66 | (42.3) | 2 | (12.5) | |

| | | | | | | |
|---|---|---|---|---|---|---|
| Close contact and use PPE above 100% of the time | 0 (0.0) | 0 (0.0) | 0 (0.0) | 0 (0.0) | 0 (0.0) | 0 (0.0) |
| Close contact and use PPE ≤ 100% of the time | 120 (58.8) | 90 (75.0) | 9 (7.5) | 5 (4.2) | 90 (57.7) | 14 (87.5) |

Abbreviations: Interquartile range (IQR), Healthcare personnel (HCP), First responders (FR), Messenger RNA (mRNA), Not applicable (N/A)

*P-values calculated using Pearson's chi-squared test or Fisher's exact test for cells with <5 observations; Kruskal Wallis non-parametric tests was used to compare median values.

† Analytic sample excludes 1,147 participants with documented SARS-CoV-2 infection before enrollment or as part of surveillance prior to the study period. Socio-demographic information was collected by self-report as part of an electronic enrollment survey.

‡ Comparison of the three vaccination groups, cohort locations for Portland, OR, Duluth, MN, Salt Lake City UT were combined compared to Phoenix, AZ, Tucson, AZ, Other, AZ, Miami, FL and Temple, TX with chi-square value of 13.1 (p-value 0.0014)

§ Comparison of any vaccination versus unvaccinated, cohort locations for Portland, OR, Duluth, MN, Salt Lake City UT were combined and compared to Phoenix, AZ, Tucson, AZ, Other, AZ, Miami, FL and Temple, TX with chi-square value of 13.0 (p-value 0.0003)

‖ Occupation categories: Primary HCP (physicians, physician assistants, nurse practitioners, dentists), Other allied HCP (nurses, therapists, technicians, medical assistants, orderlies and all others providing clinical support in inpatient or outpatient settings), first responders (FR; firefighters, law enforcement, corrections, emergency medical technicians), essential and frontline workers (EFW; workers in hospitality, delivery, and retail; teachers; all other occupations that require contact within 3 feet of the public, customers, or co-workers as a routine part of their job)

¶ For 7 participants, who did not respond to the self-report question, they were imputed as none, pending further verification.

**Each month, participants were asked about close contacts and PPE use during the past 7 days. The mean of monthly responses during the study period were calculated.

‡ ‡ Only applicable for participants indicating a potential exposure during the past 7 days.

**Table S6. Indicators of potential vaccine attenuation by participant characteristics among those with RT-PCR-confirmed SARS-CoV-2 infection**

| | All SARS-CoV-2 RT-PCR-Positives | | Viral RNA Load, Log10 Copies/mL | | | Symptom Duration | | | Days in Bed | | | Febrile CLI | | Afebrile CLI[¶] | | | RT-PCR Positive >2 weeks | | RT-PCR Positive 1 week | | |
|---|---|---|---|---|---|---|---|---|---|---|---|---|---|---|---|---|---|---|---|---|---|
| | N | (Col %) | Mean | (SD) | p-value | Mean | (SD) | p-value | Mean | (SD) | p-value | N | (Col. %) | N | (Col %) | p-value* | N | (Col. %) | N | (Col %) | p-value* |
| All participants[†] | 204 | | 3.6 | (1.7) | | 15.3 | (14.4) | | 3.2 | (5.3) | | 116 | (56.9) | 88 | (43.1) | | 135 | (66.2) | 69 | (33.8) | |
| Socio-demographic characteristics | | | | | | | | | | | | | | | | | | | | | |
| Cohort location | | | | | 0.6966 | | | 0.0029 | | | 0.1017 | | | | | 0.0025 | | | | | 0.8252 |
| Phoenix, AZ | 43 | (8.5) | 3.3 | (1.7) | | 18.2 | (11.3) | | 4.7 | (8.7) | | 24 | (20.7) | 19 | (21.6) | | 30 | (22.2) | 13 | (9.6) | |
| Tucson, AZ | 75 | (6.2) | 3.7 | (1.7) | | 16.6 | (14.6) | | 2.9 | (3.3) | | 52 | (44.8) | 23 | (26.1) | | 50 | (37.0) | 25 | (18.5) | |
| Other, AZ | 15 | (5.2) | 3.4 | (2) | | 14.2 | (14.2) | | 2.7 | (3.3) | | 9 | (7.8) | 6 | (6.8) | | 11 | (8.1) | 4 | (3.0) | |
| Miami, FL | 23 | (9.7) | 3.7 | (1.6) | | 15 | (22.4) | | 2.5 | (4.2) | | 9 | (7.8) | 14 | (15.9) | | 14 | (10.4) | 9 | (6.7) | |
| Duluth, MN | 11 | (2.2) | 3.9 | (1.6) | | 15.5 | (15.6) | | 5.2 | (8.9) | | 9 | (7.8) | 2 | (2.3) | | 9 | (6.7) | 2 | (1.5) | |
| Portland, OR | 5 | (0.8) | 3.7 | (2.3) | | 17.4 | (7) | | 4.6 | (1.7) | | 3 | (2.6) | 2 | (2.3) | | 3 | (2.2) | 2 | (1.5) | |
| Temple, TX | 18 | (5.7) | 3.6 | (2.1) | | 8.9 | (10.6) | | 1.6 | (2.3) | | 8 | (6.9) | 10 | (11.4) | | 10 | (7.4) | 8 | (5.9) | |
| Salt Lake City, UT | 14 | (3.0) | 4.3 | (1.2) | | 8.7 | (7.3) | | 1.9 | (3.2) | | 2 | (1.7) | 12 | (13.6) | | 8 | (5.9) | 6 | (4.4) | |
| Sex | | | | | 0.1489 | | | 0.8845 | | | 0.2974 | | | | | 0.7511 | | | | | 0.105 |
| Female | 111 | (4.4) | 3.5 | (1.7) | | 14.8 | (12.5) | | 3.8 | (6.4) | | 62 | (53.4) | 49 | (55.7) | | 68 | (50.4) | 43 | (31.9) | |
| Male | 93 | (6.3) | 3.8 | (1.7) | | 16 | (16.4) | | 2.5 | (3.5) | | 54 | (46.6) | 39 | (44.3) | | 67 | (49.6) | 23 | (17.0) | |
| Age(Years) | | | | | 0.5757 | | | 0.9234 | | | 0.4347 | | | | | 0.8189 | | | | | 0.7548 |
| 18-49 | 142 | (4.9) | 3.7 | (1.7) | | 14.6 | (12.4) | | 3.3 | (5.9) | | 80 | (69.0) | 62 | (70.5) | | 93 | (68.9) | 49 | (36.3) | |
| ≥50 | 62 | (5.5) | 3.5 | (1.7) | | 17 | (18.1) | | 3 | (3.8) | | 36 | (31.0) | 26 | (29.5) | | 42 | (31.1) | 20 | (14.8) | |
| Race | | | | | 0.6455 | | | 0.8889 | | | 0.9613 | | | | | 0.6063 | | | | | 0.062 |
| White | 178 | (5.1) | 3.6 | (1.6) | | 15 | (13.9) | | 3,3 | (5.6) | | 100 | (86.2) | 78 | (88.6) | | 122 | (90.4) | 56 | (41.5) | |
| Other | 26 | (5.1) | 3.7 | (1.7) | | 17.3 | (17.4) | | 2.6 | (2.7) | | 16 | (13.8) | 10 | (11.4) | | 13 | (9.6) | 13 | (9.6) | |
| Ethnicity | | | | | 0.7638 | | | 0.7932 | | | 0.0900 | | | | | 0.068 | | | | | 0.1264 |
| Hispanic/Latinx | 60 | (8.5) | 3.7 | (1.6) | | 16 | (15) | | 3.9 | (5.4) | | 40 | (34.5) | 20 | (22.7) | | 35 | (25.9) | 25 | (18.5) | |
| Other | 144 | (4.4) | 3.6 | (1.7) | | 15.1 | (14.2) | | 2.9 | (5.3) | | 76 | (65.5) | 68 | (77.3) | | 100 | (74.1) | 44 | (32.6) | |

| | | | | | | | | | | | | | |
|---|---|---|---|---|---|---|---|---|---|---|---|---|---|
| Occupation‡ | | | 0.0254 | | 0.9637 | | 0.4043 | | | 0.7307 | | | 0.6604 |
| Primary HCP | 16 ( 2.0 ) | 3.5 ( 1.7 ) | | 13.8 ( 10.9 ) | | 1.3 ( 1.3 ) | | 8 ( 6.9 ) | 8 ( 9.1 ) | | 11 ( 8.1 ) | 5 ( 3.7 ) | |
| Nurses and other allied HCP | 66 ( 4.9 ) | 3.3 ( 1.8 ) | | 14.9 ( 11.9 ) | | 3.5 ( 4.8 ) | | 37 ( 31.9 ) | 29 ( 33.0 ) | | 40 ( 29.6 ) | 26 ( 19.3 ) | |
| First Responders | 73 ( 9.2 ) | 4.1 ( 1.5 ) | | 15.2 ( 15.2 ) | | 2.9 ( 4.2 ) | | 45 ( 38.8 ) | 28 ( 31.8 ) | | 49 ( 36.3 ) | 24 ( 17.8 ) | |
| Essential and other frontline | 49 ( 4.5 ) | 3.4 ( 1.8 ) | | 16.6 ( 17.2 ) | | 3.8 ( 7.6 ) | | 26 ( 22.4 ) | 23 ( 26.1 ) | | 35 ( 25.9 ) | 14 ( 10.4 ) | |
| Chronic Condition | | | 0.3464 | | 0.6684 | | 0.5598 | | | 0.2295 | | | 0.1133 |
| None § | 139 ( 5.1 ) | 3.5 ( 1.8 ) | | 15.4 ( 14.2 ) | | 3.1 ( 5.7 ) | | 83 ( 71.6 ) | 56 ( 63.6 ) | | 87 ( 64.4 ) | 52 ( 38.5 ) | |
| 1 or more | 65 ( 5.1 ) | 3.7 ( 1.7 ) | | 15.1 ( 14.9 ) | | 3.3 ( 4 ) | | 33 ( 28.4 ) | 32 ( 36.4 ) | | 48 ( 35.6 ) | 17 ( 12.6 ) | |

Abbreviations: Interquartile range (IQR), Healthcare personnel (HCP), COVID-19-like illness (CLI)

*P-values calculated using Pearson's chi-squared test or Fisher's exact test for cells with <5 observations; Kruskal Wallis non-parametric tests was used to compare median values.

† Analytic sample excludes 1,147 participants with documented SARS-CoV-2 infection before enrollment or as part of surveillance prior to the study period. Socio-demographic information was collected by self-report as part of an electronic enrollment survey.

‡ Occupation categories: Primary HCP (physicians, physician assistants, nurse practitioners, dentists), Other allied HCP (nurses, therapists, technicians, medical assistants, orderlies and all others providing clinical support in inpatient or outpatient settings), first responders (FR; firefighters, law enforcement, corrections, emergency medical technicians), essential and frontline workers (EFW; workers in hospitality, delivery, and retail; teachers; all other occupations that require contact within 3 feet of the public, customers, or co-workers as a routine part of their job)

§ For 7 participants, who did not respond to the self-report question, they were imputed as none, pending further verification.

¶Afebrile defined as anyone who didn't report fever or chills in surveys

**<u>Figure_S1. Percent SARS-CoV-2 positive of all tested in local counties and dates of PCR-confirmed infections by site location (panels A-H)</u>**

A

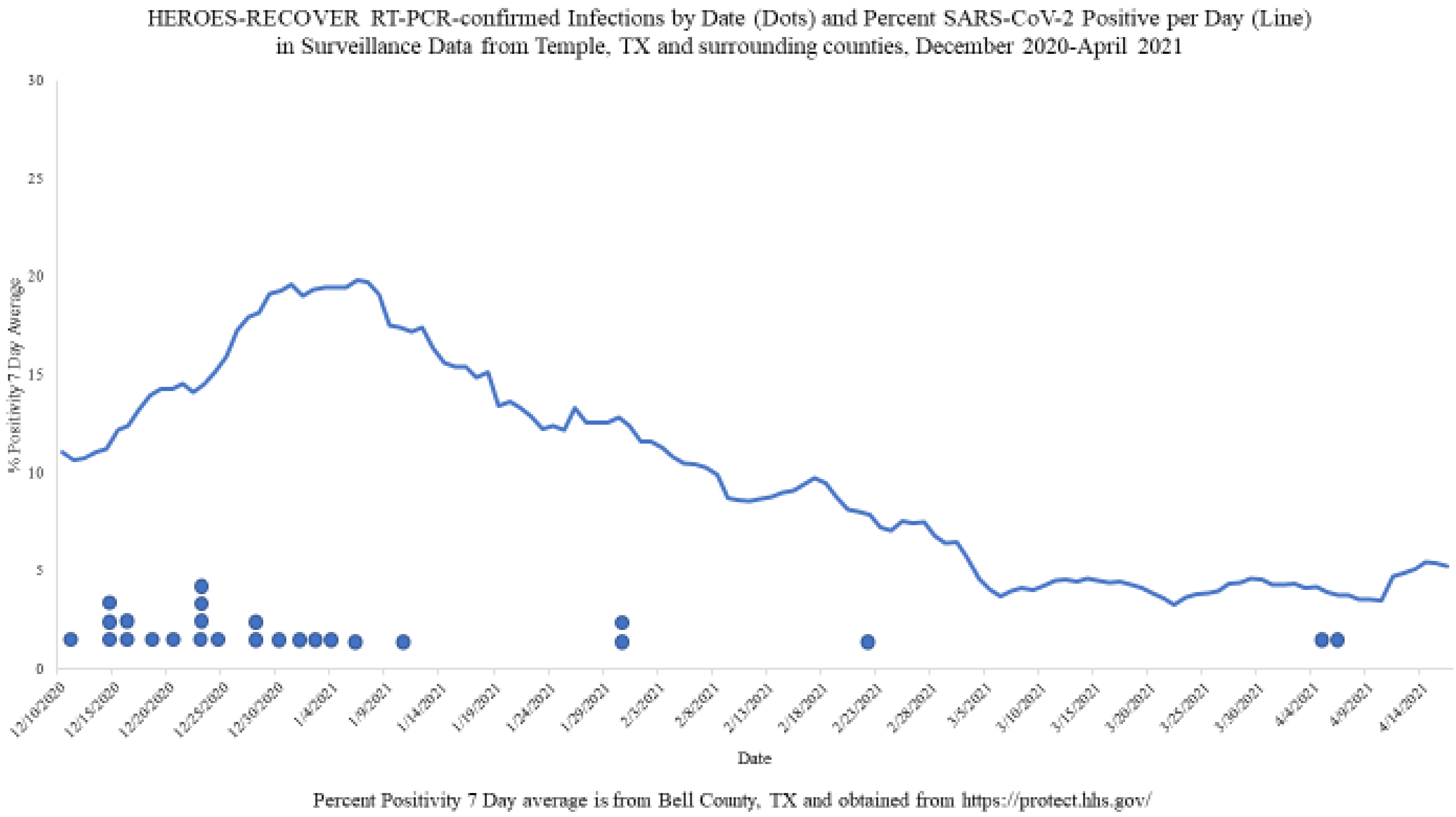

B

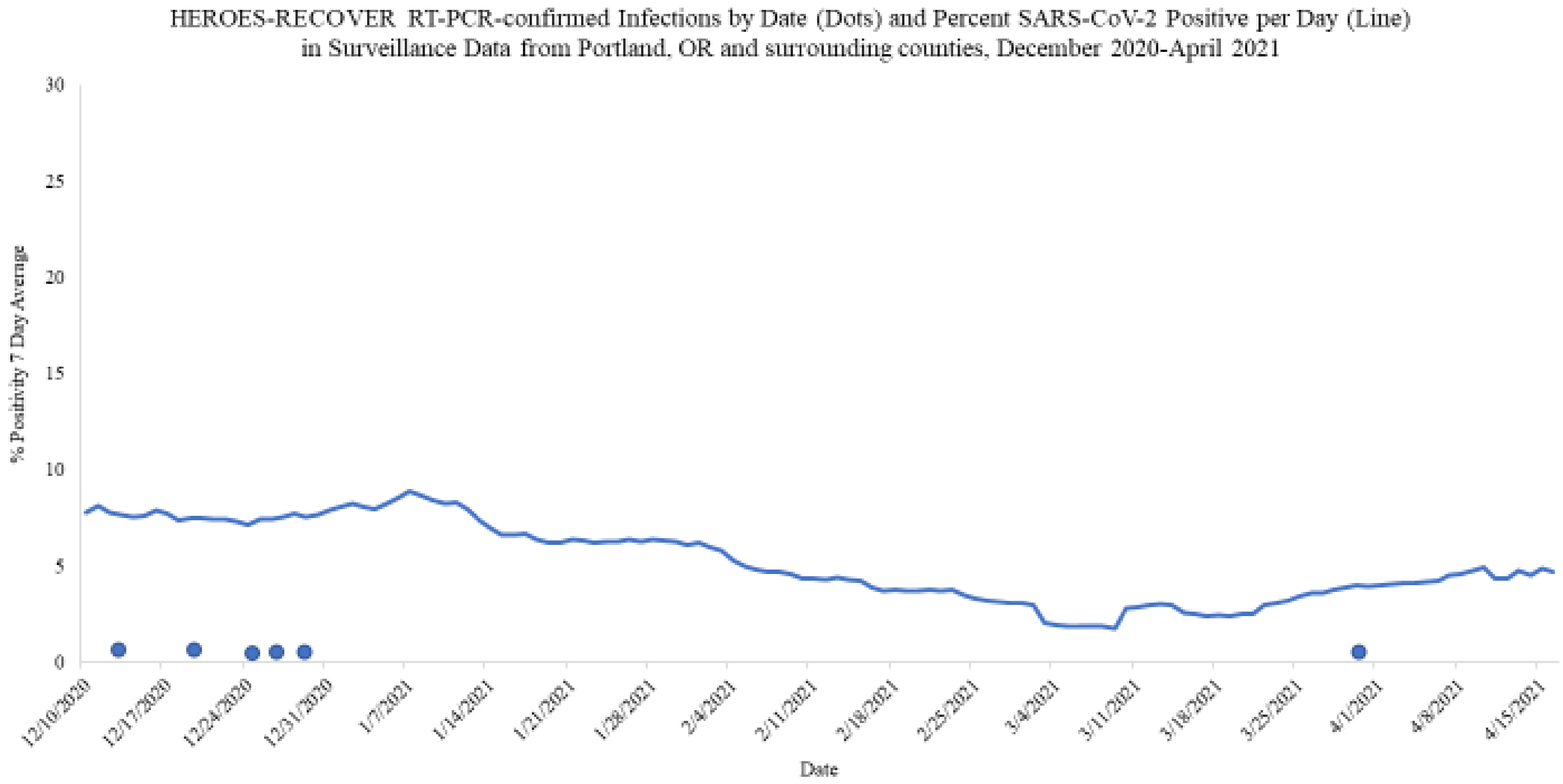

Percent Positivity 7 Day average is from Clackamas, Multnomah and Washington Counties, OR and obtained from https://protect.hhs.gov/

C

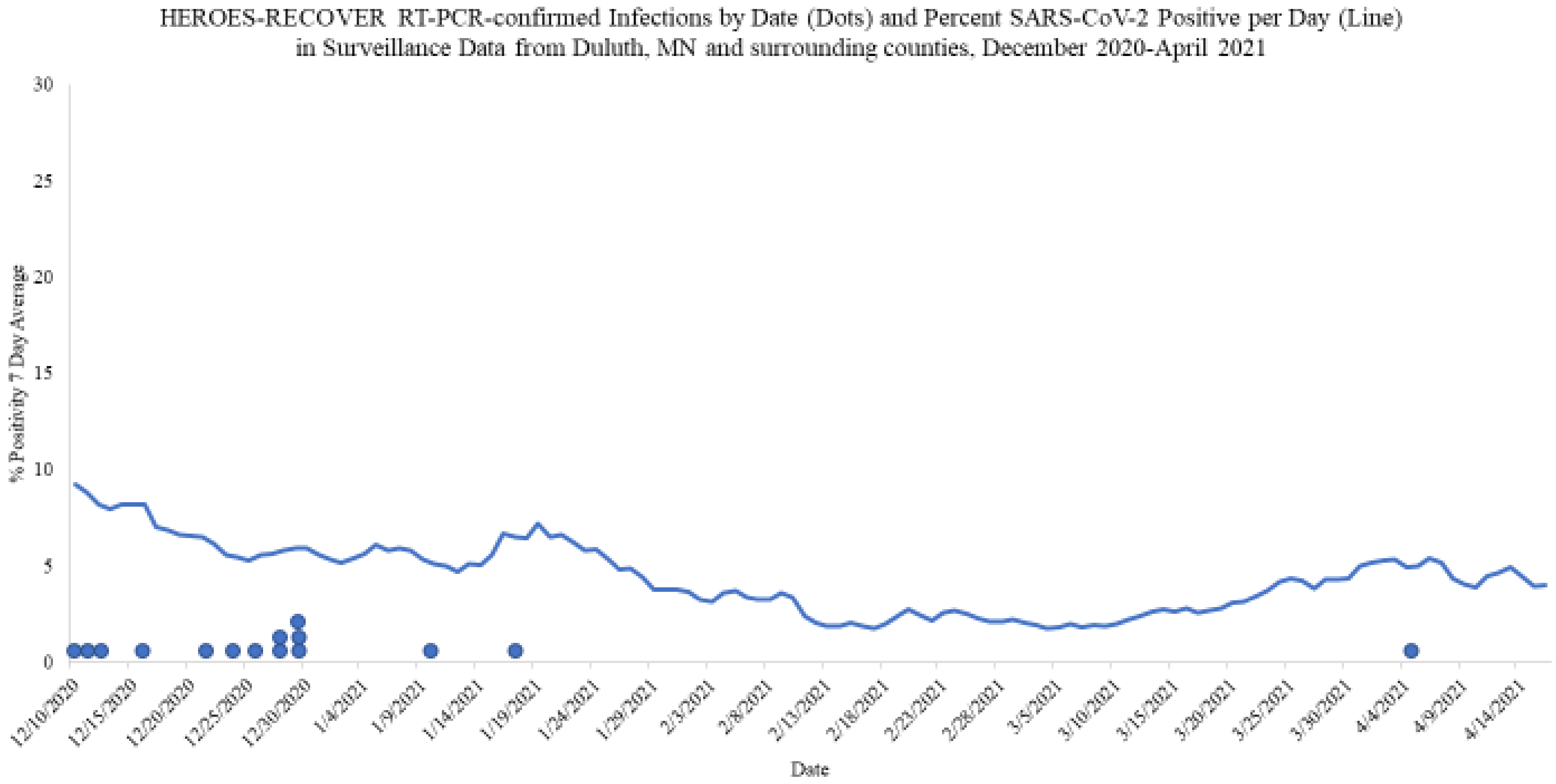

Percent Positivity 7 Day average is from Carlton, Douglas, Lake and St. Louis Counties, MN as well as Ashland and Bayfield Counties, WI. Data was obtained from https://protect.hhs.gov

D

HEROES-RECOVER RT-PCR-confirmed Infections by Date (Dots) and Percent SARS-CoV-2 Positive per Day (Line) in Surveillance Data from Tucson, AZ and surrounding counties, December 2020-April 2021

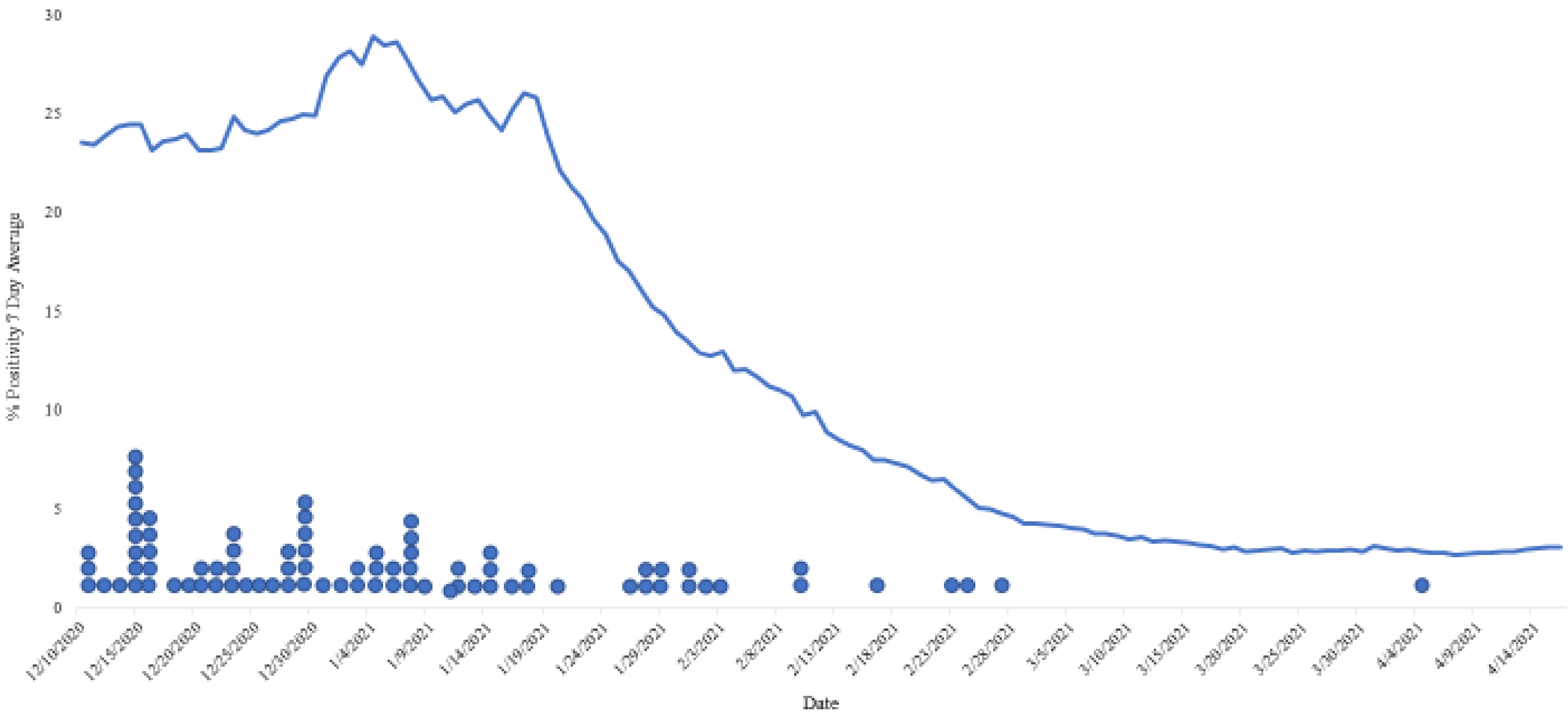

Percent Positivity 7 Day average is from Pima County, AZ and obtained from https://protect.hhs.gov

E

HEROES-RECOVER RT-PCR-confirmed Infections by Date (Dots) and Percent SARS-CoV-2 Positive per Day (Line) in Surveillance Data from Salt Lake City, UT and surrounding counties, December 2020-April 2021

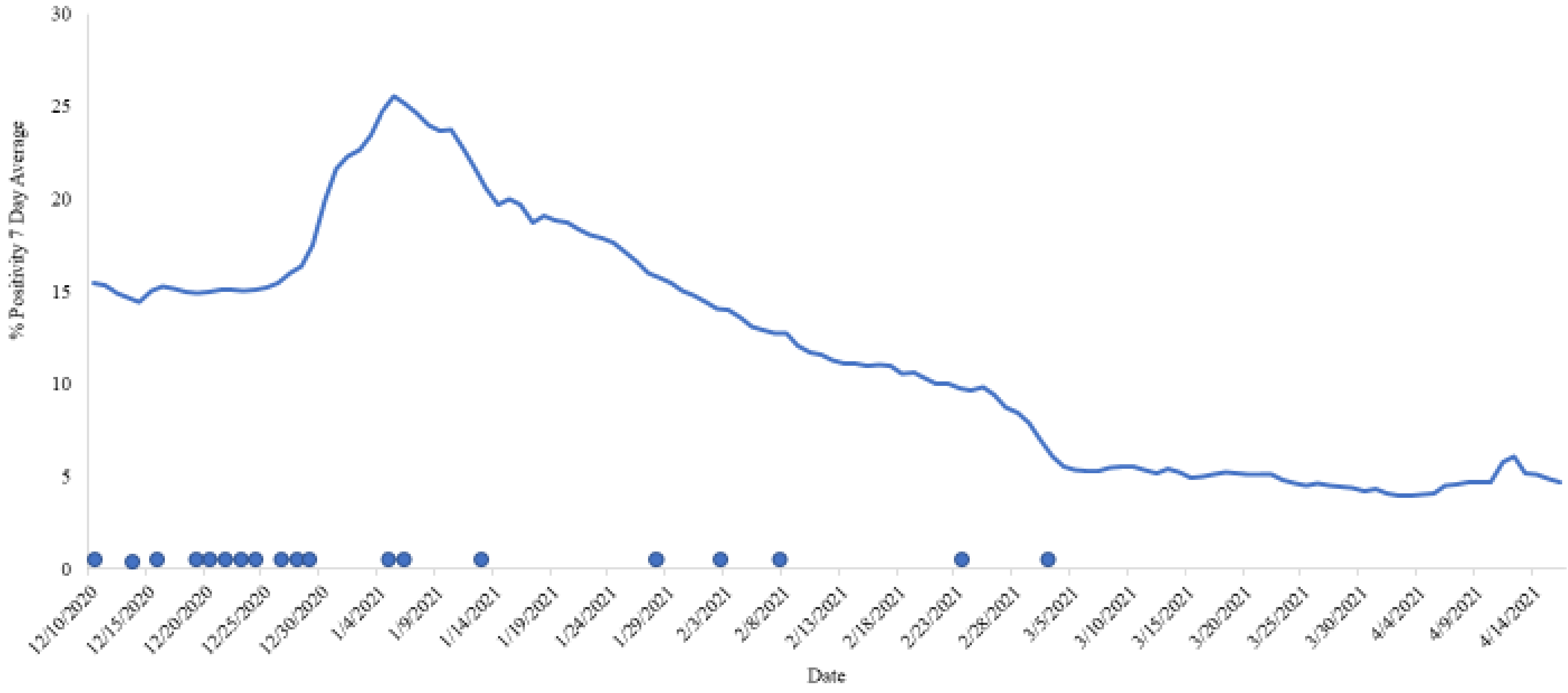

Percent Positivity 7 Day average is from Davis, Salt Lake, Summit, Tooele, Utah and Weber Counties, UT and obtained from https://protect.lhs.gov

F

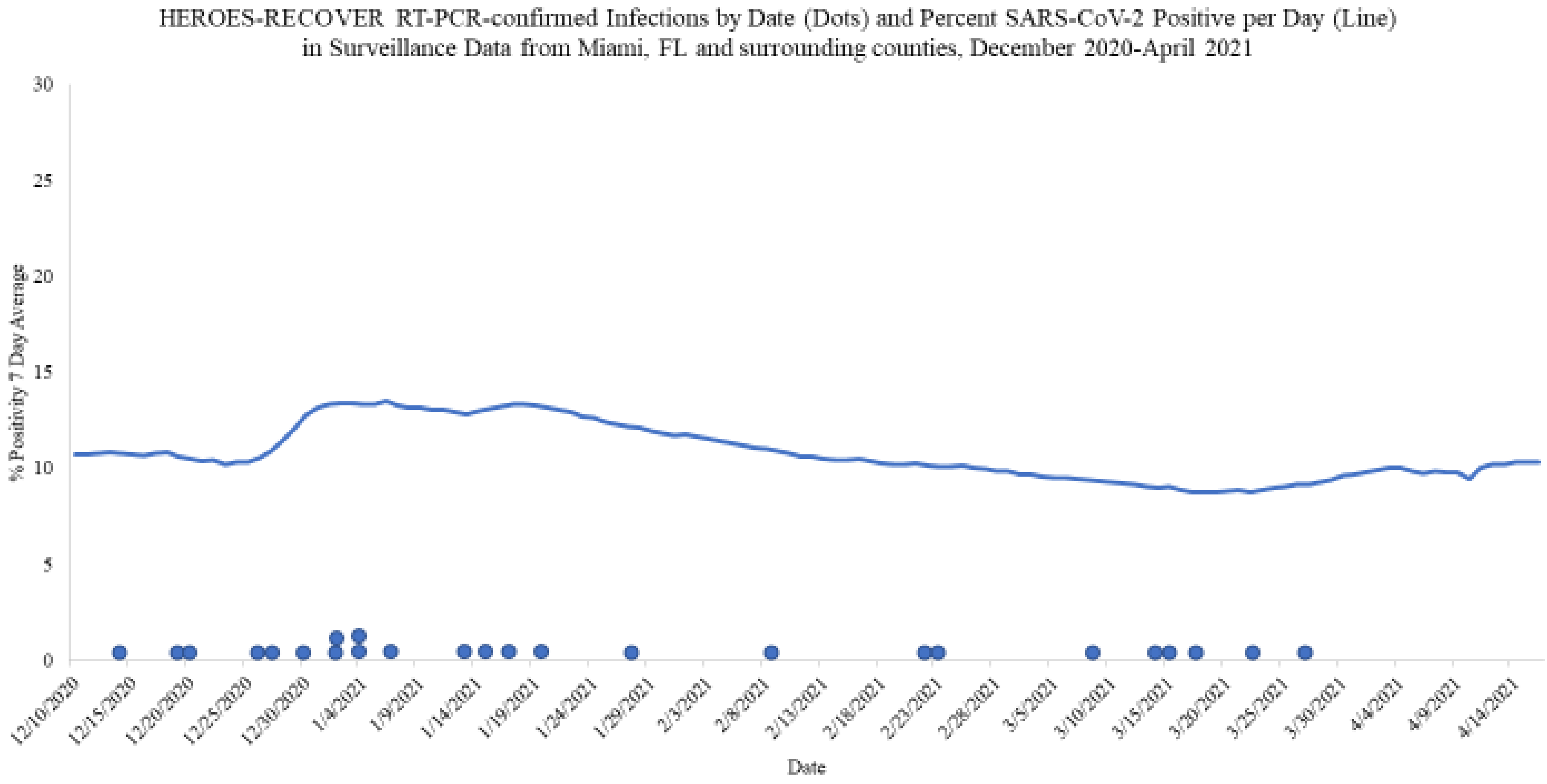

Percent Positivity 7 Day average is from Broward, Miami-Dade and Palm Beach Counties, FL and obtained from https://protect.hhs.gov

G

HEROES-RECOVER RT-PCR-confirmed Infections by Date (Dots) and Percent SARS-CoV-2 Positive per Day (Line) in Surveillance Data from Phoenix, AZ and surrounding counties, December 2020-April 2021

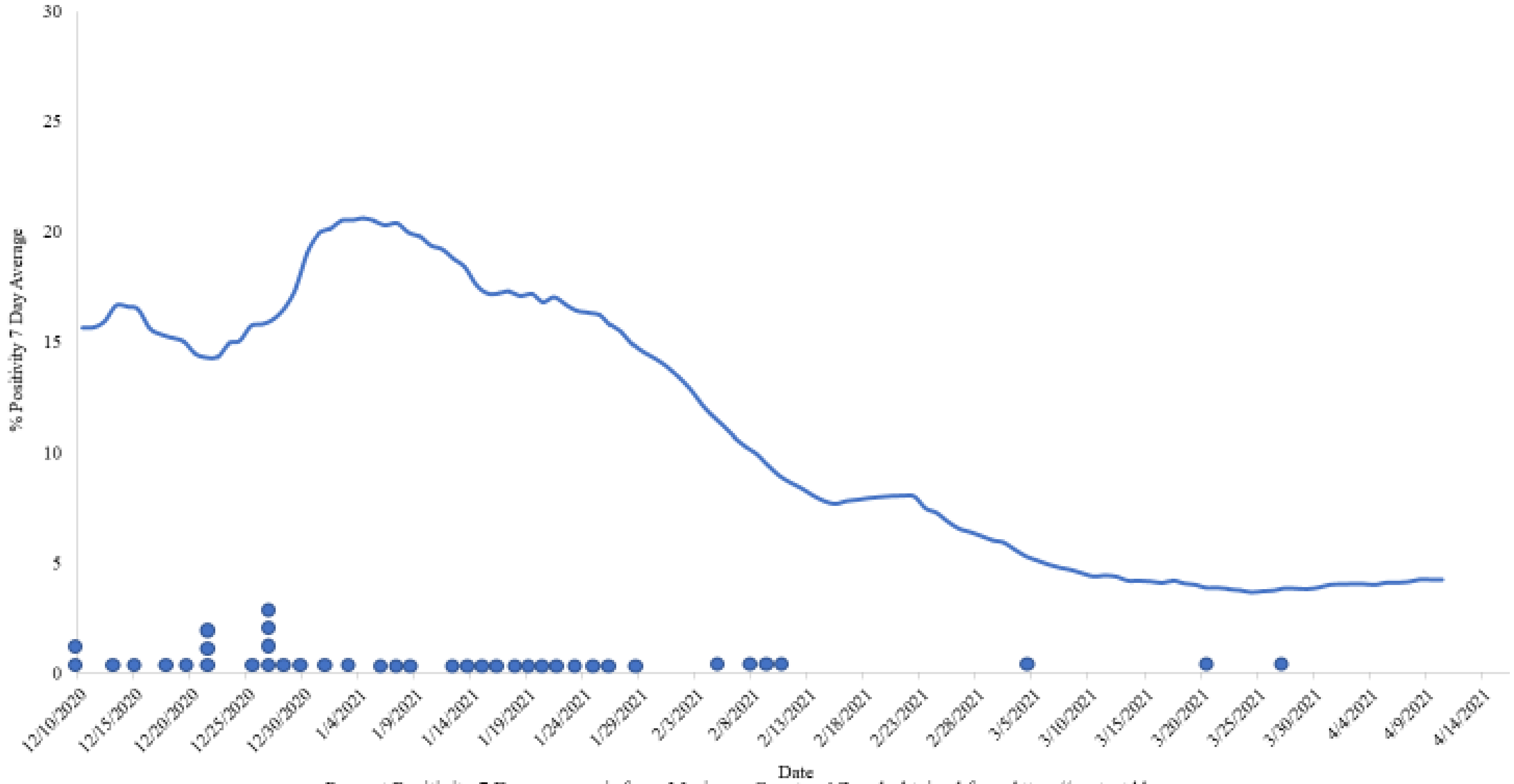

Percent Positivity 7 Day average is from Maricopa County, AZ and obtained from https://protect.hhs.gov

H

HEROES-RECOVER RT-PCR-confirmed Infections by Date (Dots) and Percent SARS-CoV-2 Positive per Day (Line) in Surveillance Data from Other AZ, December 2020-April 2021

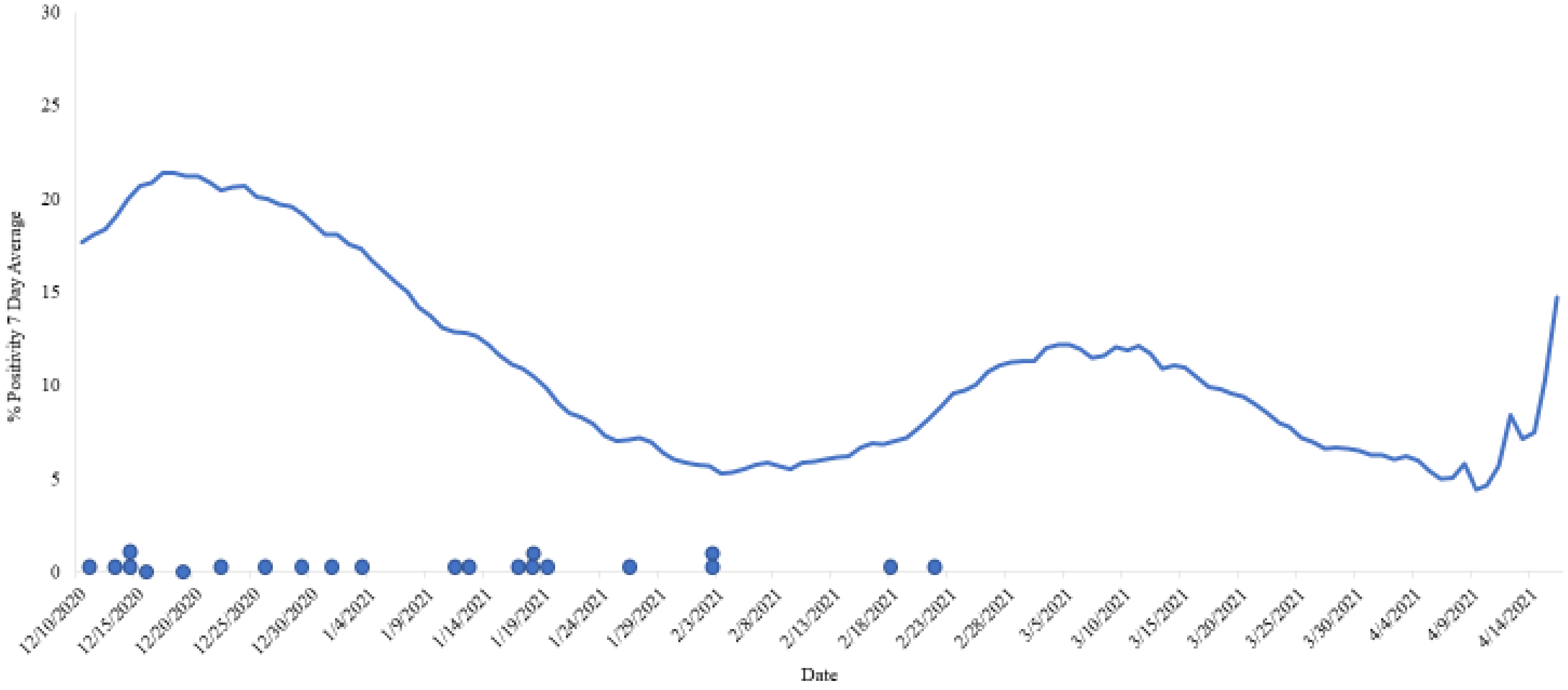

Percent Positivity 7 Day average is from Apache, Cochise, Coconino, Gila, Graham, Greenlee, La Paz, Mohave, Navajo, Pinal, Santa Cruz, Yavapai, and Yuma Counties, A[illegible] obtained from https://protect.hhs.gov

**Figure S2. CONSORT diagram of HEROES-RECOVER prospective cohort participants**

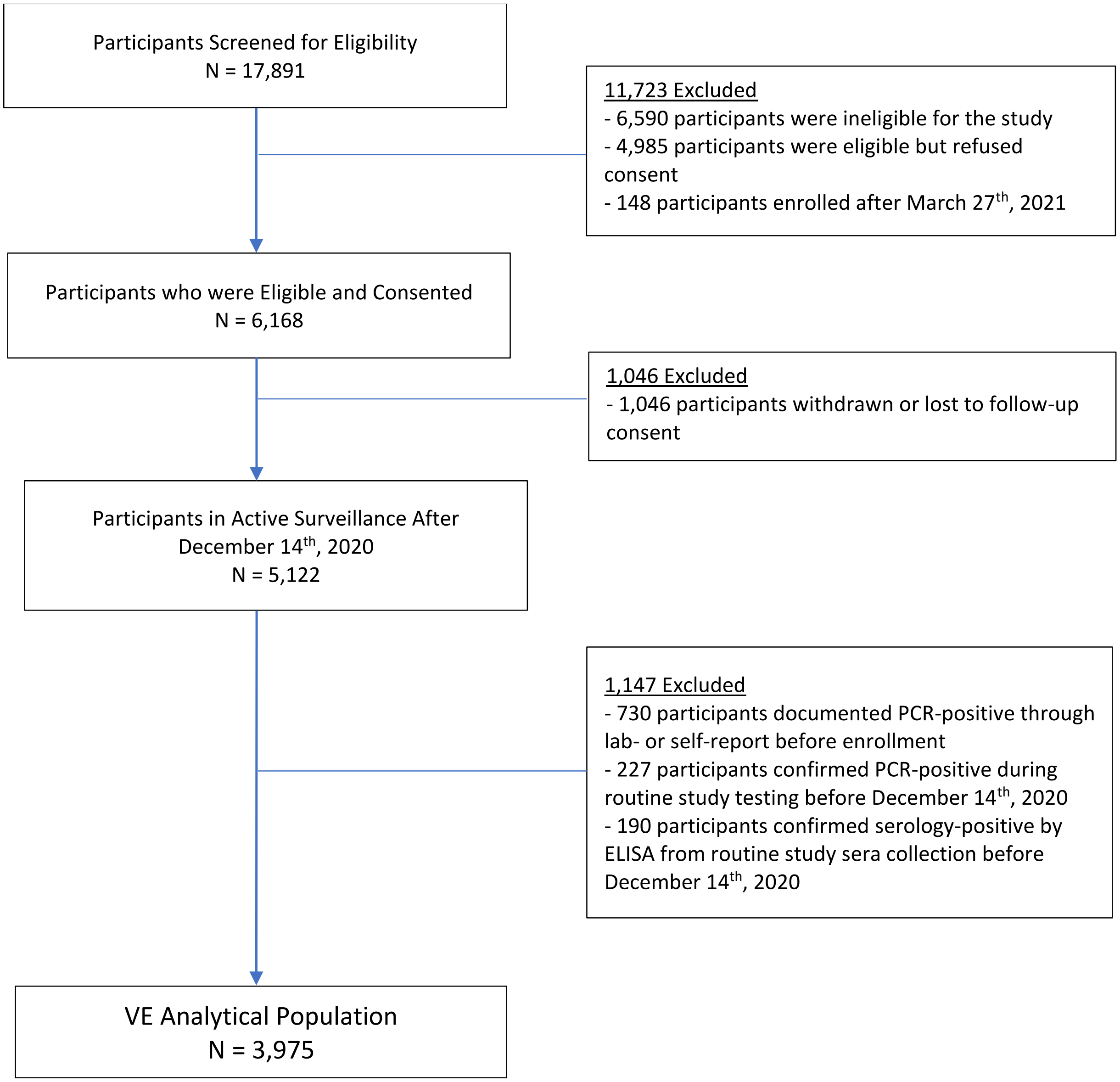

**Figure_S3. Standardized mean differences of covariates between unvaccinated and vaccinated participants with receipt of at least one dose before and after inverse propensity of treatment weighting.**

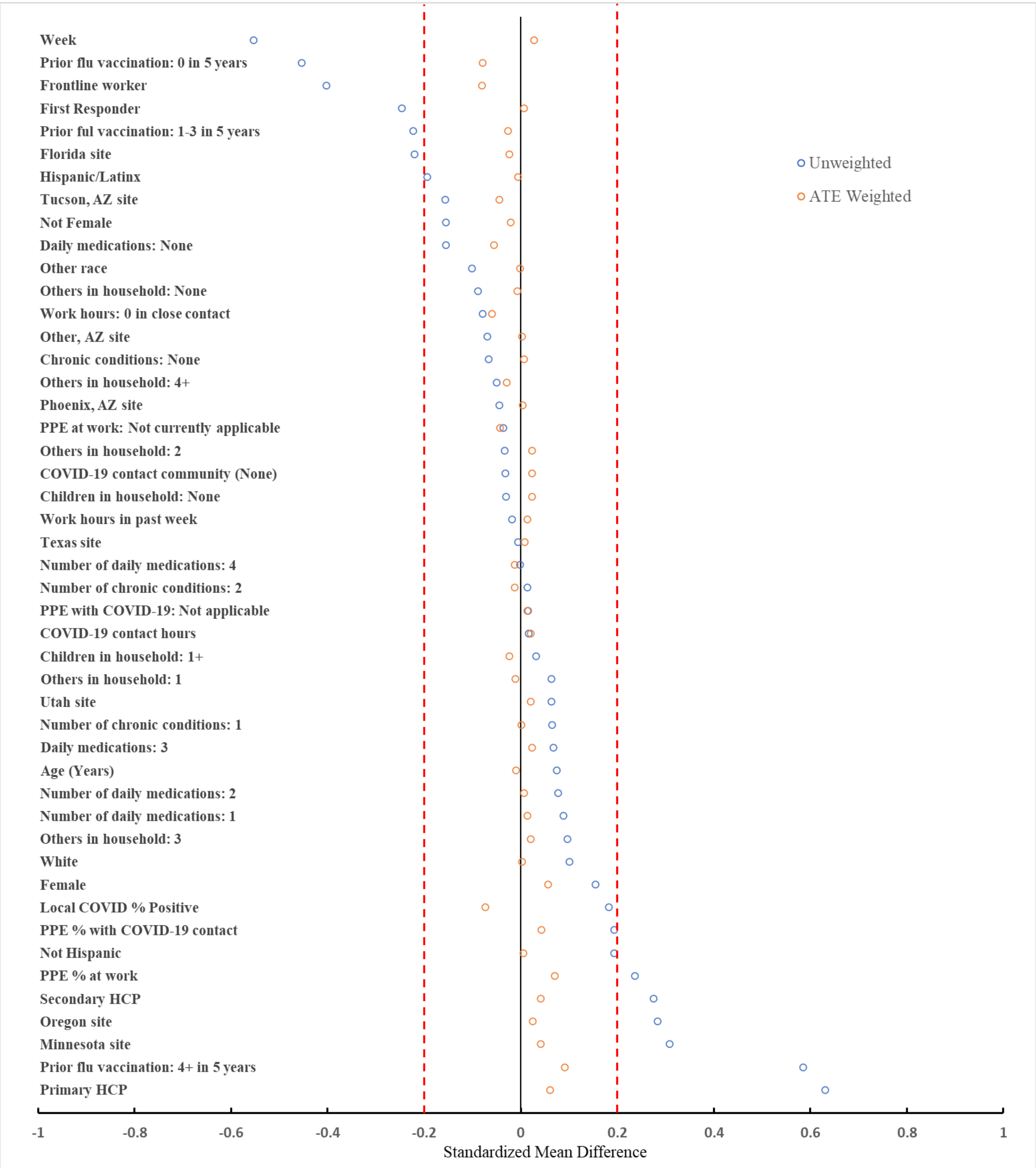

Legend: Negative differences indicate groups that are less likely to be vaccinated and positive differences indicate those more likely to be vaccinated. Absolute standard mean differences of less than 0.2 are considered well balanced. The largest difference after ATE weighting was 0.09.

Abbreviation: Average treatment effect weighted (ATE)

**Figure_S4. SARS-CoV-2 viral RNA load among all participants with RT-PCR-confirmed infection after receipt of one dose of mRNA vaccine (n=50), by day post dose 1**

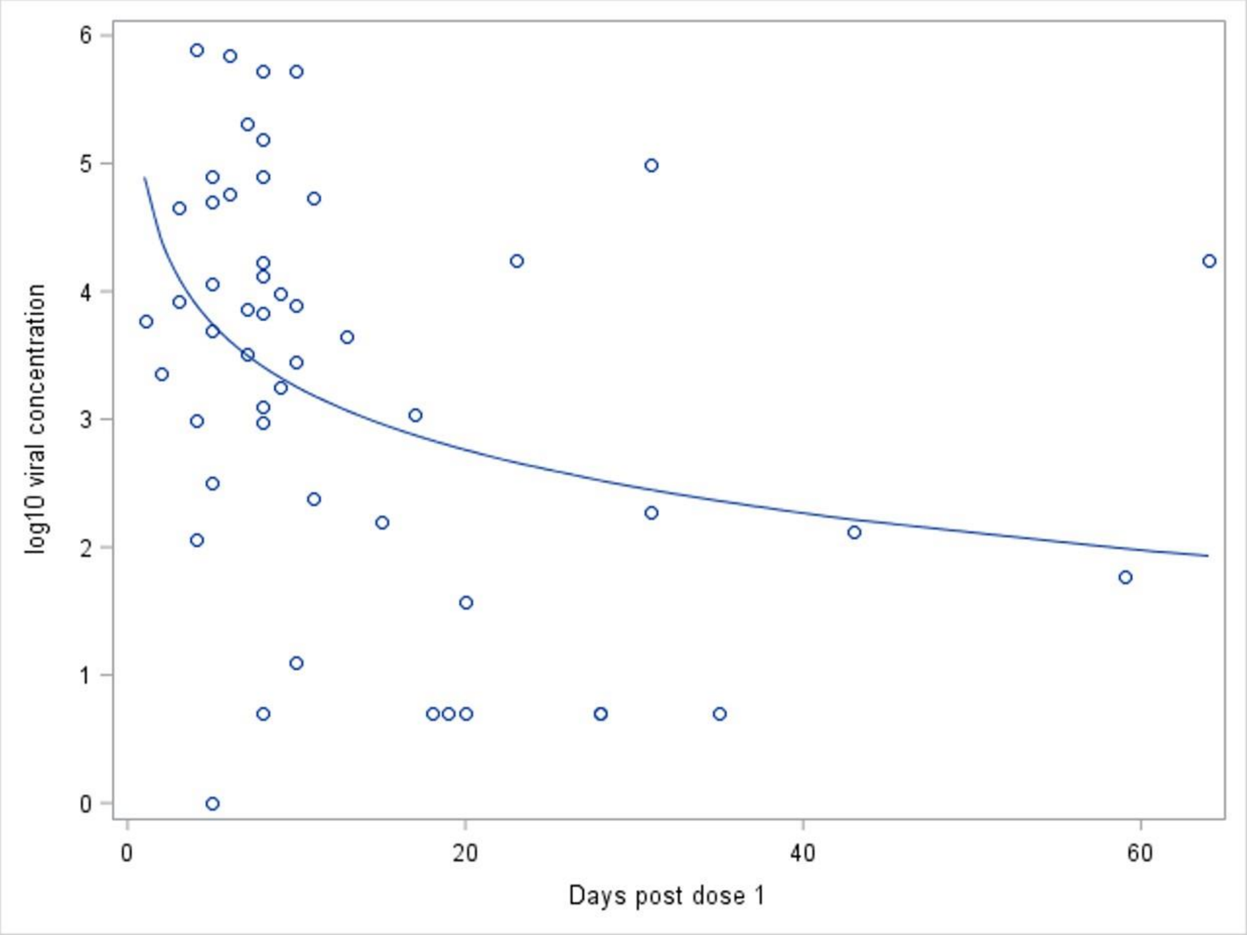
